# Parenclitic network mapping predicts survival in critically ill patients with sepsis

**DOI:** 10.1101/2025.01.16.25320643

**Authors:** Emily Ito, Tope Oyelade, Matthew Wikner, Jinyuan Liu, Watjana Lilaonitkul, Ali R Mani

## Abstract

Sepsis is a complex disease involving multiple organ systems. A network physiology approach to sepsis may reveal collective system behaviours and intrinsic organ interactions. However, mapping functional connectivity for individual patients has been challenging due to the lack of analytical methods for evaluating physiological networks using routine clinical and laboratory data. This study explored the use of parenclitic network mapping to assess organ connectivity and predict sepsis outcomes based on routine laboratory data. Data from 162 sepsis patients meeting Sepsis-3 criteria were retrospectively analysed from the MIMIC-III database, focusing on 48-hour deterioration and 30-day survival. Fifteen physiological variables representing organ systems were used to construct organ network connectivity through correlation analysis. Correlation analysis identified 7 organ interactions linked to 30-day survival and 9 related to 48-hour deterioration. Parenclitic network analysis was used to measure deviations in individual patients’ correlations between organ systems from the reference physiological interactions observed in survivors. Parenclitic deviations in the pH-bicarbonate axis (hazard ratio=2.081, p<0.001) and pH-lactate axis (hazard ratio=2.773, p=0.024) significantly predicted 30-day mortality, independent of the Sequential Organ Failure Assessment (SOFA) score and ventilation status. However, parenclitic deviations did not predict 48-hour deterioration. This study highlights the potential of parenclitic network mapping to provide insights into sepsis pathophysiology and differences in organ system connectivity between survivors and non-survivors independent of sepsis severity and mechanical ventilation status. Integrating physiological network mapping with current prognostic tools may enhance personalized sepsis management and outcome prediction in clinical settings.

**New and Noteworthy:** This study employs an integrative network analysis approach to investigate sepsis, aiming to leverage routine laboratory data for prognostication and to enhance understanding of the pathophysiology of sepsis. Using parenclitic network mapping, we assessed individual organ connectivity patterns that distinguish between survivors and non-survivors. Our findings demonstrate that physiological network mapping in sepsis offers prognostic insights independent of disease severity and mechanical ventilation status in critically ill patients with sepsis.

## Introduction

Sepsis is a complex life-threatening disorder due to dysregulated host response to systemic infection that leads to multiorgan failure (Singer *et al*., 2016). Global burden of sepsis is substantial, with around 48.9 million estimated worldwide cases and 11 million deaths annually (Fleischmann *et al*., 2016, Rudd *et al*., 2020). The body’s response to systemic infection in sepsis is highly *dynamic*. Immune homeostasis which is essential for orchestrating effective response against infection is dysregulated in sepsis, causing hemodynamic and metabolic changes that lead to organ dysfunction (Salomão *et al*., 2019). While improved intensive care units (ICU) management has increased survival rates in the early hyperinflammatory phase of sepsis, the persistence of the disease can result in a shift from immune resistance to immune tolerance (Delano and Ward, 2016). Patients who develop immunosuppression suffer from long-term mortality due to impaired capacity to eradicate both the primary pathogen and nosocomial infections (Denstaedt, Singer and Standiford, 2018). This makes sepsis a complex disease as both hyperinflammation and immunosuppression play an important role in the progression of sepsis and its survival (Hotchkiss, Monneret and Payen, 2013). Physiological organs response to systemic inflammation is diverse and depends on metabolic state and energetic trade-offs which eventually may lead to deterioration, organ dysfunction or disease tolerance (Ganeshan *et al*., 2019). Moreover, factors such as age, infection origin, and comorbidities also influence the sepsis trajectory, contributing to disease heterogeneity (Denstaedt, Singer and Standiford, 2018).

Over the past decades, numerous attempts have been made to develop effective treatments, but most clinical trials have failed due to challenges posed by the complex pathophysiology of sepsis and patient heterogeneity (Marshall, 2014). There is strong evidence suggesting that early identification and treatment of at-risk sepsis population (e.g. early identification of deterioration) improves patient outcome (Seymour *et al*., 2017). Current methods for evaluating sepsis like Sequential Organ Failure Assessment (SOFA) are based on linear models of sepsis pathophysiology, where the severity of organ dysfunction is estimated by treating each organ as an independent entity and summing the level of dysfunction across all organs (Figure 1.a.) (Vincent *et al*., 1996). However, these methods have a limited ability to identify early sepsis, potentially due to the simplistic nature of the linear models which can overlook the complex interplay between organ systems that is generated in response to systemic infection during sepsis (Godin and Buchman, 1996; Moorman, Lake and Ivanov, 2016; Quinten *et al*., 2018).

**Figure 1.**
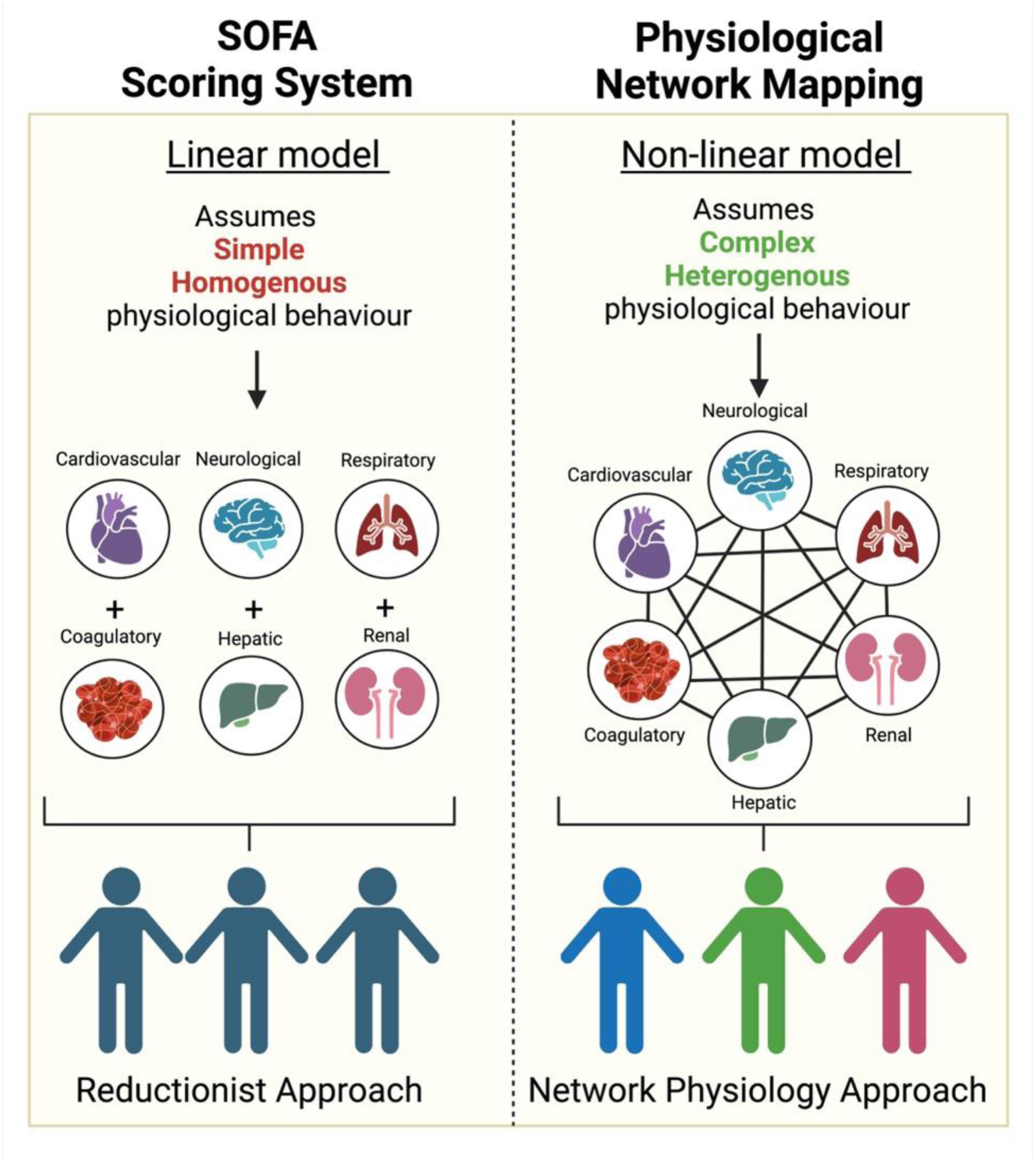
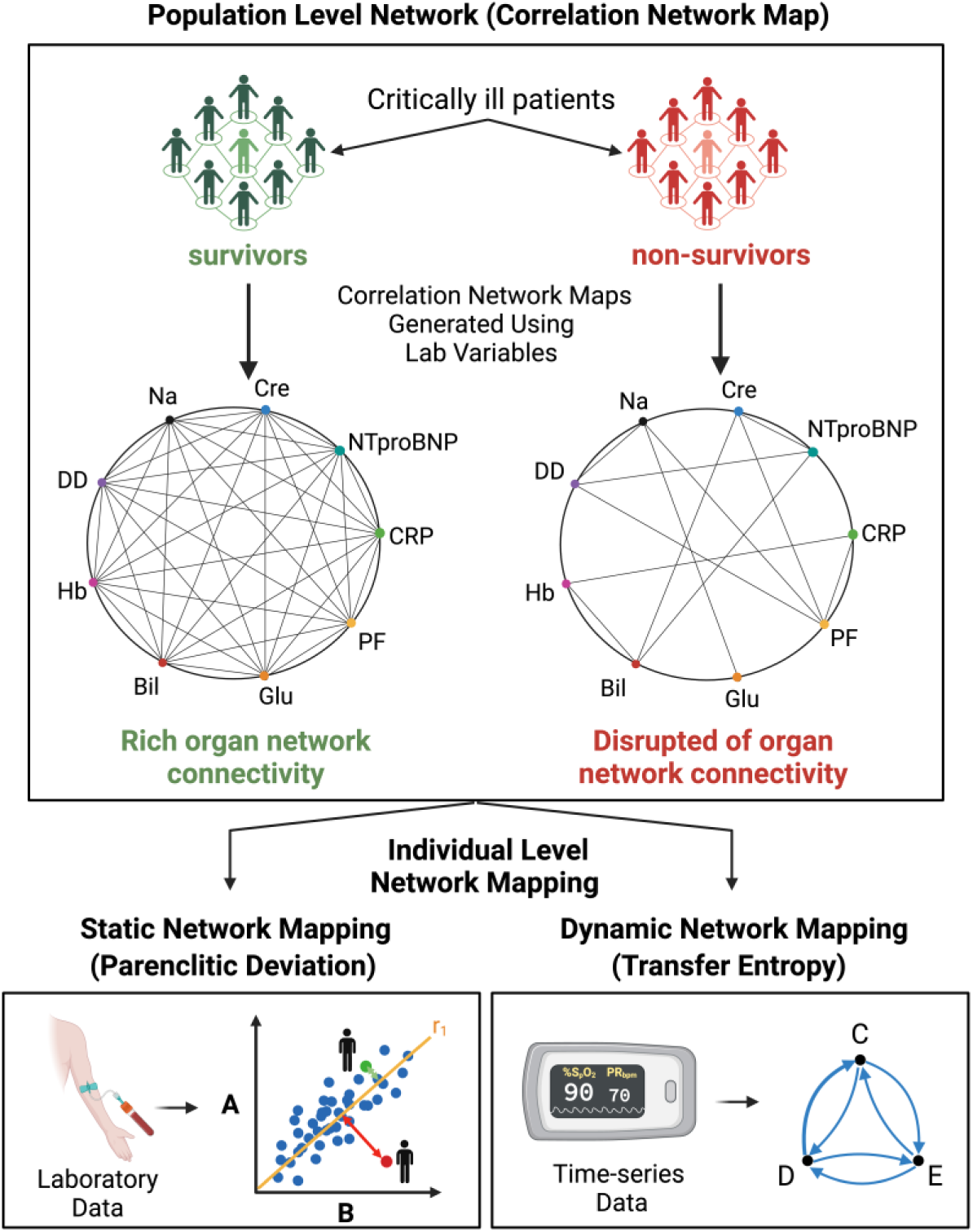
**a.** Diagram illustrating the differences between linear and non-linear models of sepsis pathophysiology. (Created with Biorender.com) **b.** Diagram illustrating different network physiological techniques used for population level (A) and individual level (B) analysis of organ network connectivity (Created with Biorender.com)

Network physiology is an emerging field that studies complex interactions within the physiological system (Bashan *et al*., 2012). Unlike SOFA which takes a linear approach, network physiology approach considers the interaction between physiological components, making it suitable for investigating physiological behaviours in complex diseases like sepsis (Figure 1.a.) (Godin and Buchman, 1996; Asada *et al*., 2016).

Numerous studies have highlighted the critical role of organ system connectivity in health and disease (Asada *et al*., 2016; Bhogal and Mani, 2017; Jiang *et al*., 2021; Zhang *et al*., 2022; Oyelade *et al*., 2023; Morandotti *et al*., 2025). A pioneering report by Asada et al. investigating organ network connectivity in critically ill patients has yielded some promising findings (Asada *et al*., 2016). The study examined the physiological network in patients by selecting physiological variables from the blood test that represent different organ systems and analysing the correlation amongst these variables. They showed that in critically ill patients with matched severity of organ dysfunction (i.e., SOFA), non-survivors had much less significant correlations between physiological variables compared to survivors. These findings lead to two important conclusions: 1) organ network disruption is associated with poor patient outcomes and 2) studying organ interactions may yield independent information regarding patient outcomes, in addition to linear models like SOFA. There is growing evidence showing that sepsis is associated with the uncoupling of physiological compensatory mechanisms and reduced organ system engagement (Godin and Buchman, 1996; Gholami *et al*., 2012; Papaioannou *et al*., 2012; Gheorghita *et al*., 2022). Therefore, studying organ connectivity in sepsis might yield important information about the patient outcomes.

The correlation network mapping approach introduced by Asada et al. is an effective technique for analysing physiological network interactions, but it is limited to population level analyses. In contrast, techniques such as parenclitic deviation (PD) and transfer entropy (TE) investigates organ network connectivity in individual patients using static (i.e., laboratory variables) and dynamic data (i.e., physiological time-series such as heart rate) respectively (Figure 1.b.) (Schreiber, 2000; Zanin *et al*., 2014). PD evaluates physiological interactions by looking at how individual patient data deviate from the reference population characteristics (i.e., in survivors) (Zhang *et al*., 2022). Meanwhile, TE measures organ connectivity by computing the bidirectional information transfer between physiological systems (Schreiber, 2000).

A recent report by Morandotti et al. employed TE analysis to study the cardio-respiratory network in septic patients using publicly available data obtained from the MIMIC-III database (Johnson *et al*., 2016; Morandotti *et al*., 2025). The study demonstrated that evaluation of organ connectivity in sepsis patients using TE could predict 30-day survival and 48-hour deterioration, independent of SOFA and ventilation status. While TE shows potential as novel tool for detecting early sepsis deterioration, its clinical application may be challenging due to the need for noise-free time-series data which can be hard to obtain in intensive care units, particularly in resource-limited settings (Berntson and Stowell, 1998; Lee and Jung, 2018).

One major advantage of PD computation is that it can be computed using a single measurement from routine laboratory test (i.e., routine blood test), requiring minimal patient involvement. Parenclitic analysis has demonstrated positive results in predicting disease outcomes for numerous conditions: such as acute liver failure, cirrhosis, and ovarian and prostate cancer (Whitwell *et al*., 2018; Zhang *et al*., 2022; Morandotti *et al*., 2025; Oyelade, Moore and Mani, 2024). However, network mapping with PD has never been applied in sepsis research.

This research builds on the previous work by Morandotti et al. and investigates the same sepsis population from the MIMIC-III database (Morandotti *et al*., 2025). The primary aim of this study was to investigate whether evaluation of organ network connectivity through parenclitic analysis of routinely collected physiological data can predict 48-hour deterioration and 30-day mortality in sepsis patients.

## Materials and Methods

This retrospective cohort study was performed using electronic medical records from critically ill sepsis patients admitted to Beth Israel Deaconess Medical Center in Boston, Massachusetts, obtained through the Medical Information Mart for Intensive Care (MIMIC-III) database (Johnson *et al*., 2016). An overview of the study design is illustrated in Figure 2.a.

**Figure 2.**
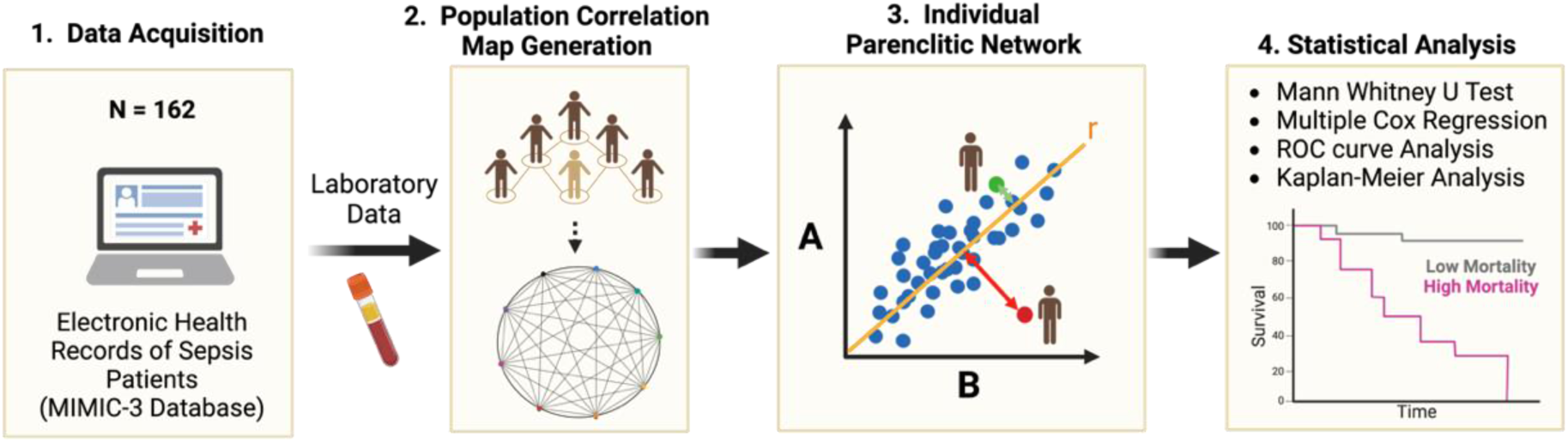
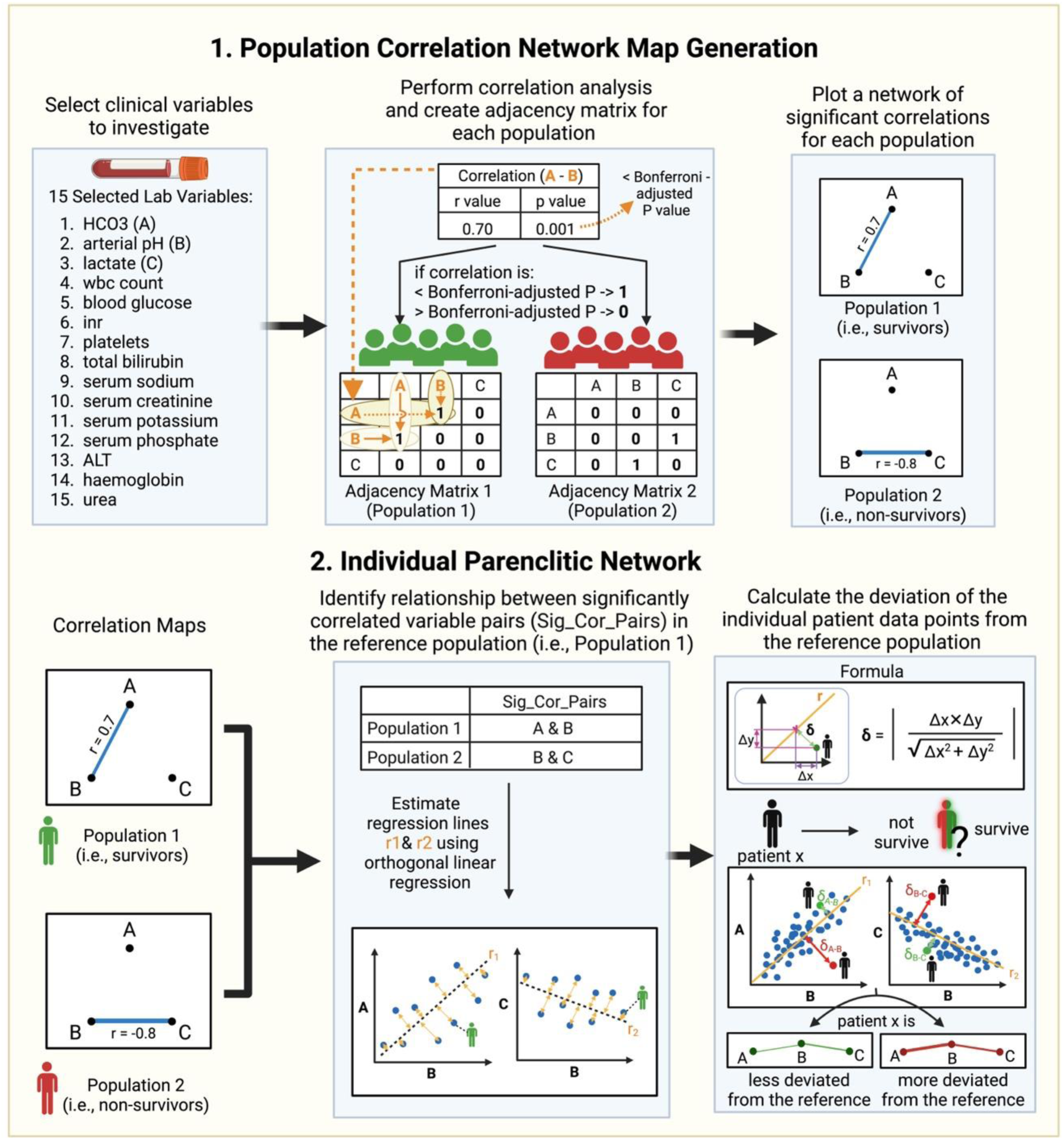
**a.** An overview of the research design. (Original diagram created with Biorender.com) **b.** Process behind parenclitic network mapping. 1) Population correlation map generation. 2) Computation of individual parenclitic network for significant physiological correlations identified from population correlation maps. “r” represents the correlation coefficient for regression line of significantly correlated physiological variables (i.e., A & B) within the reference population (i.e., survivors). “*δ*;” represents the parenclitic deviation (PD). “Δ” represents the difference between the reference population and individual patient data. All PD computation was conducted using in-house code developed in MATLAB. (Original diagram created with Biorender.com)

### Ethics Statement

Patient information from the MIMIC-III database was deidentified in compliance with Health Insurance Portability and Accountability Act (HIPAA) standards. The MIMIC project was approved by the Institutional Review Boards of Beth Israel Deaconess Medical Center and the MIT (IRB protocol nos. 2001P001699/14 and 0403000206, respectively). Prior to the study, the authors involved in data processing completed a mandatory online ethics training at MIT and were accredited (IDs: 10304625 and 48067739).

### Patient Data Extraction

Structure query language (SQL) code was used to extract patient data from the MIMIC-III database. The study population included patients above 18 years old with a sepsis diagnosis who had a single record of ICU admission. Sepsis patients were selected based on Sepsis 3 criteria: acute increase in SOFA score ≥ 2 points and a suspected infection (Seymour *et al*., 2016; Singer *et al*., 2016).

This paper builds on the work by Morandotti et al. and studied the same patient cohort (N = 164) (Morandotti *et al*., 2025). Instead of using waveform data for dynamic network mapping as done by Morandotti et al., this study focused on investigating the laboratory data of the sepsis patients for static network mapping. Patients from this cohort were selected for the present study if they met the above inclusion criteria and had matching descriptive clinical data, waveform recordings, and laboratory results. Two patients were excluded because their waveform data and laboratory results were recorded at different times. As a result, 162 patients were included in the correlation network mapping and the parenclitic analysis. A flowchart detailing the complete data extraction process is shown in Supplementary material Figure S1.

The clinical data extracted include the ventilation status, SOFA score, 30-day survival follow-up, 30-day survival status, and 48-hour deterioration outcome. The SOFA score was calculated on two occasions: first, on the earliest day when both the laboratory and waveform data were available, and second, after 48-hours. Deterioration was defined as an increase in SOFA by at least 2 points at 48-hours. The details of SOFA calculation and the definition of deterioration is described elsewhere (Morandotti *et al*., 2025).

For the laboratory data, 15 physiological variables representing different organ systems were retrieved: including serum phosphate, arterial pH, urea, haemoglobin, lactate, white blood cell count (WBC), serum sodium, international normalised ratio (INR), platelets, total bilirubin, blood glucose, serum creatinine, alanine transaminase (ALT), bicarbonate (HCO3), and serum potassium. The mean level of variables measured within the first 24 hours of hospital admission was used for the parenclitic analysis. All laboratory data come from routine blood test. The arterial pH has been measured directly using arterial blood gas (ABG) analyser and the HCO3 levels were obtained separately from the basic metabolic panel. The pH and the bicarbonate levels were not estimated using Henderson-Hasselbalch equation, to ensure that the correlation observed between pH and HCO3 is purely physiological and not due to the mathematical relationship.

### Correlation Network Map Generation in a Sepsis Population

Correlation network map was generated to investigate organ connectivity at the population level (Figure 2.b.). 162 patients were separated according to their 30-day survival and 48-hour deterioration outcome. Correlations between 15 physiological variables were analysed as described by Asada et al. (Asada *et al*., 2016). To account for the missingness in the data, Pearson’s correlation analysis was conducted with a pair-matching technique to omit patient data from analysis whenever either of the two variables being investigated was missing (Tan, Montagnese and Mani, 2020). Correlation between two variables was considered significant if the significance level was less than the Bonferroni-adjusted p-value (p≤0.00047619). The results from correlation analysis were used to form an adjacency matrix, where “0” indicate lack of significant correlation and “1” represents significant correlation between two physiological variables (**Figure 2.b.**). Correlation networks were visualised for 30-day survival and 48-hour deterioration by using the adjacency matrix to plot a graph. Nodes represent the physiological variables and edges show a significant correlation between two variables. The edge thickness was weighted according to the strength of Pearson’s correlation coefficient of individual correlations to illustrate the relative strength of each correlation within the network. All codes for generating the correlation network map were developed in MATLAB build R2024a (MATLAB, 2024).

### Parenclitic Network Mapping of Individual Patients

Significant correlations identified from the population correlation network maps for 30-day survival and 48-hour deterioration were used to calculate parenclitic deviations (PD). Parenclitic analysis was conducted for individual patients (N =162) by investigating how the relationship between physiological variables in each patient deviated from the general trend in the reference population, which consisted of the survivor cohort for 30-day-survival and non-deteriorated patient group for 48-hour deterioration (Zhang *et al*., 2022; Oyelade *et al*., 2023). For each pair of significant correlations between the physiological variables, its general trend in the reference population was identified by performing a regression analysis (Figure 2.b.). The orthogonal linear regression (OLS) was chosen for the regression line estimation over the classical least square (CLS) method because OLS accounts for the errors in the measurement of both dependent and independent variables, while CLS assumes measurement errors only occur in the dependent variable (Keleş, 2018). Following the estimation of the regression line representing the trend in reference population, PD was computed by measuring the orthogonal distance between each patient’s datapoint and the regression line using the equation shown in Figure 2.b. (Zhang *et al*., 2022). All the PDs were computed using an algorithm built in MATLAB build R2024a (MATLAB, 2024). The code developed by the author is publicly available on GitHub: https://github.com/emilyito/Parenclitic-Deviation-OLR.

### Statistical Analysis

All the statistical analysis was conducted using SPSS Statistics 29 (Corp., 2023). Data are reported as mean ± standard deviation and the significance level was set at 0.05 for statistical tests unless stated otherwise. Mann-Whitney U test was performed to compare differences in PDs and SOFA scores between survivors and non-survivors for 30-day survival and deteriorated and non-deteriorated patients for 48-hour deterioration. Multivariate Cox regression was performed to investigate whether the computed PD values significantly predict 30-day survival and 48-hour deterioration, independent of SOFA score and ventilation status. Since PDs look at the relationship between two variables (i.e., PD(pH-lactate)) and each variable could independently influence the disease outcome, the mean values of the analysed physiological variables (i.e., such as pH and lactate for PD(pH-lactate)) were also included as covariates. PD values used in the multivariate Cox regression were normalised using Z-transformation prior to analysis. Receiver Operating Characteristic (ROC) curve analysis was performed for PD axes that significantly predicted 30-day survival. Area Under the Curve (AUC) from the ROC curve analysis was obtained to assess and compare the diagnostic performance between PDs and SOFA. Youden index was used as a cut-off with the optimum balance of sensitivity and specificity for classifying patients into predicted survivor and non-survivor groups. Sepsis patients classified into predicted survivor and non-survivor group were used to generate the Kaplan-Meier Curve for 30-day survival. The Log-Rank test was used to compare the survival times between the two groups.

## Results

### General Characteristics of the Study Population

Out of 162 patients included in the study, 30 patients deteriorated at 48-hour follow-up, and 33 patients have passed away within 30 days of ICU admission. Sepsis patients with poor disease outcomes including non-survivors and deteriorated patients were older, had a higher SOFA score, and had a greater likelihood of having a positive ventilation status compared to survivors and non-deteriorated patients (**Table 3.a**.). The number of patients with positive ventilation status was significantly different for both 30-day survival and 48-hour deterioration (p<0.001). Significant differences in age (p=0.008) and SOFA score (p<0.001) were only found for 30-day survival. For the physiological variables, phosphate, urea, lactate, and creatinine levels were significantly elevated, and bicarbonate and arterial pH were significantly reduced in non-survivors and deteriorated patients compared to survivors and non-deteriorated patients. The white blood cell count (p<0.045) and glucose level (p<0.019) was only significantly elevated in non-survivors for 30-day survival. Conversely, alanine transaminase (p<0.043) was only significantly elevated in deteriorated patients for 48-hour deterioration.

**Table 3.**
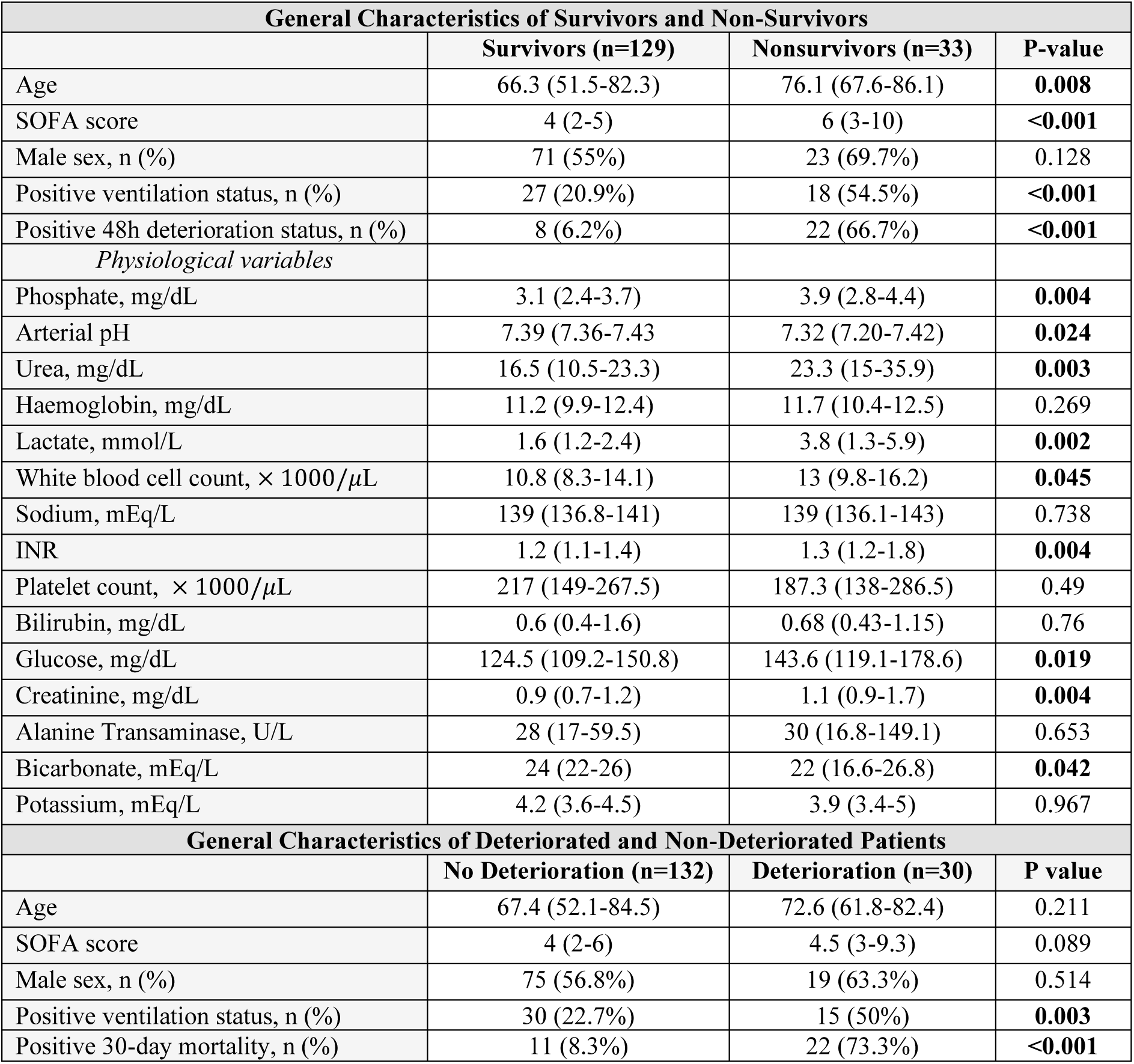
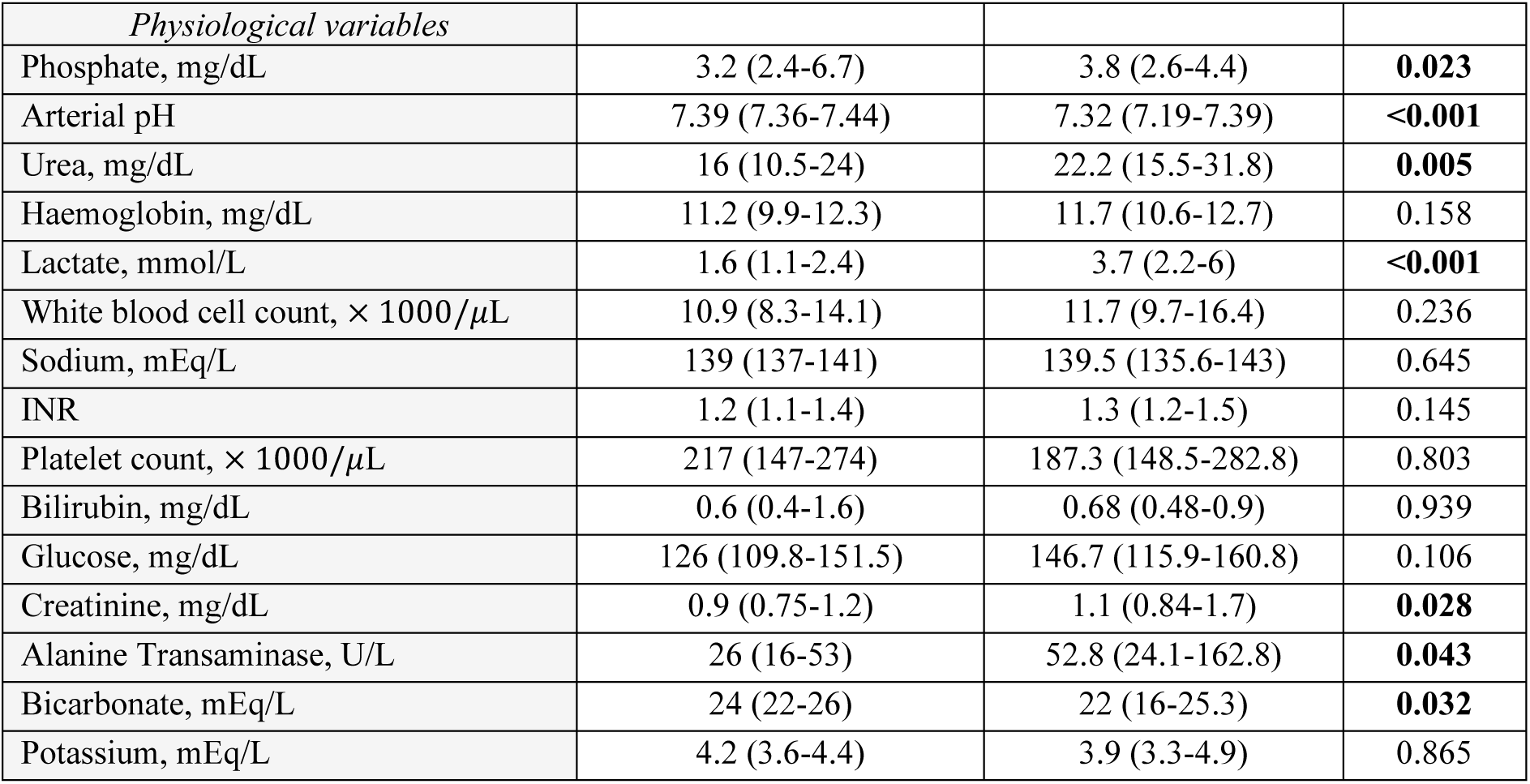
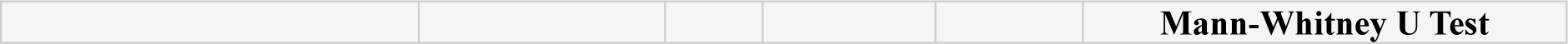
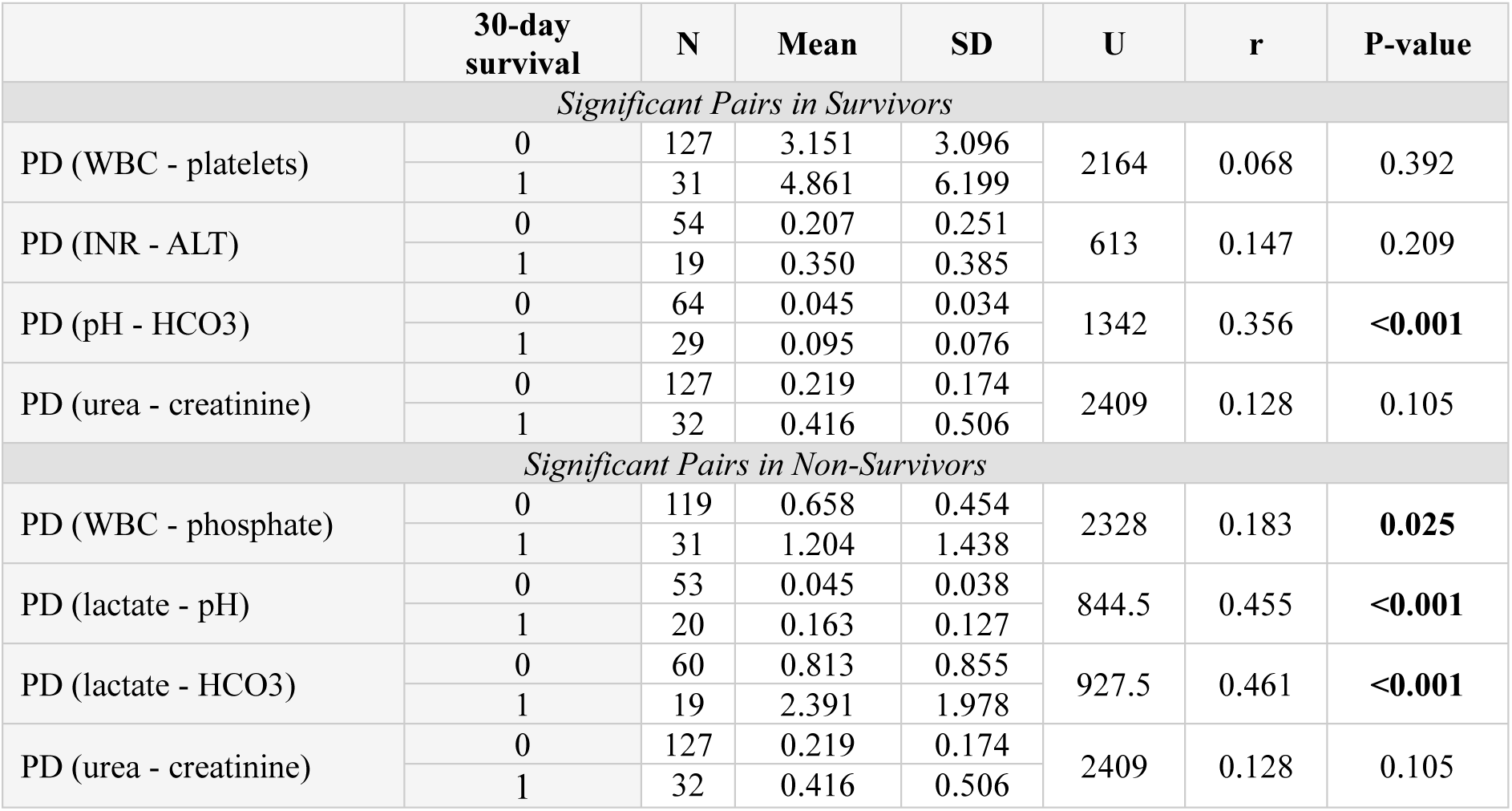
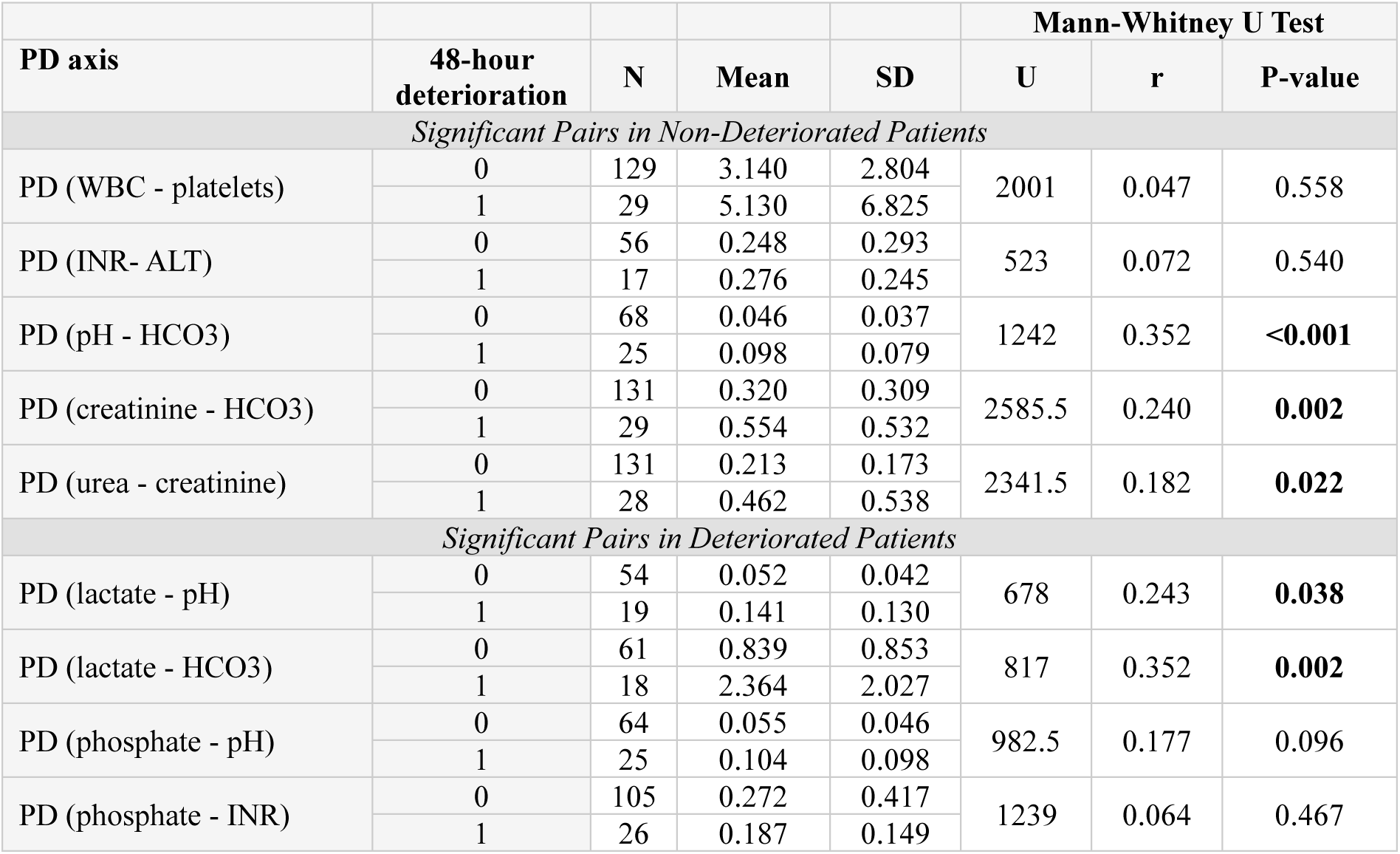
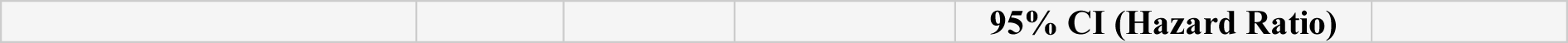
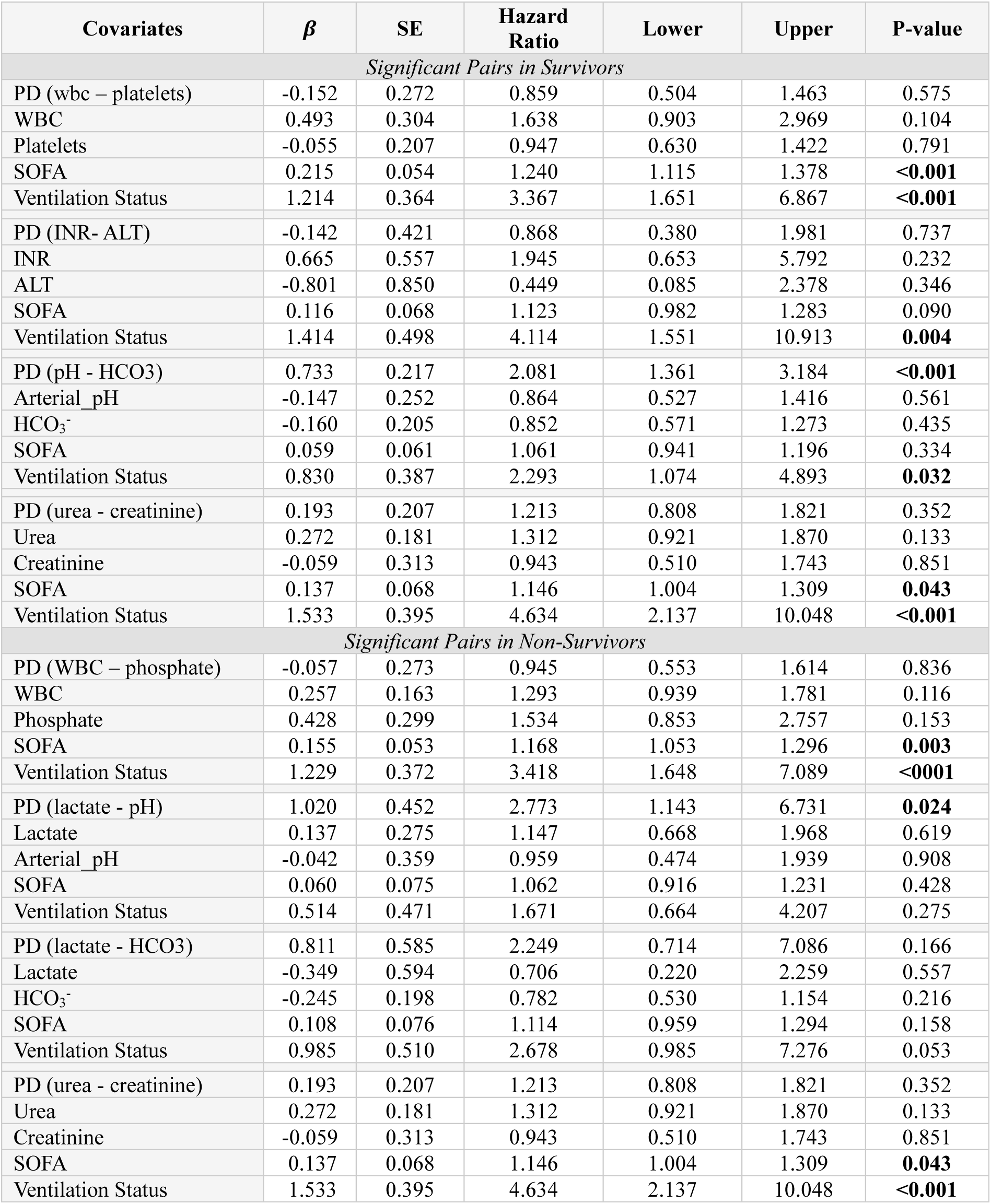
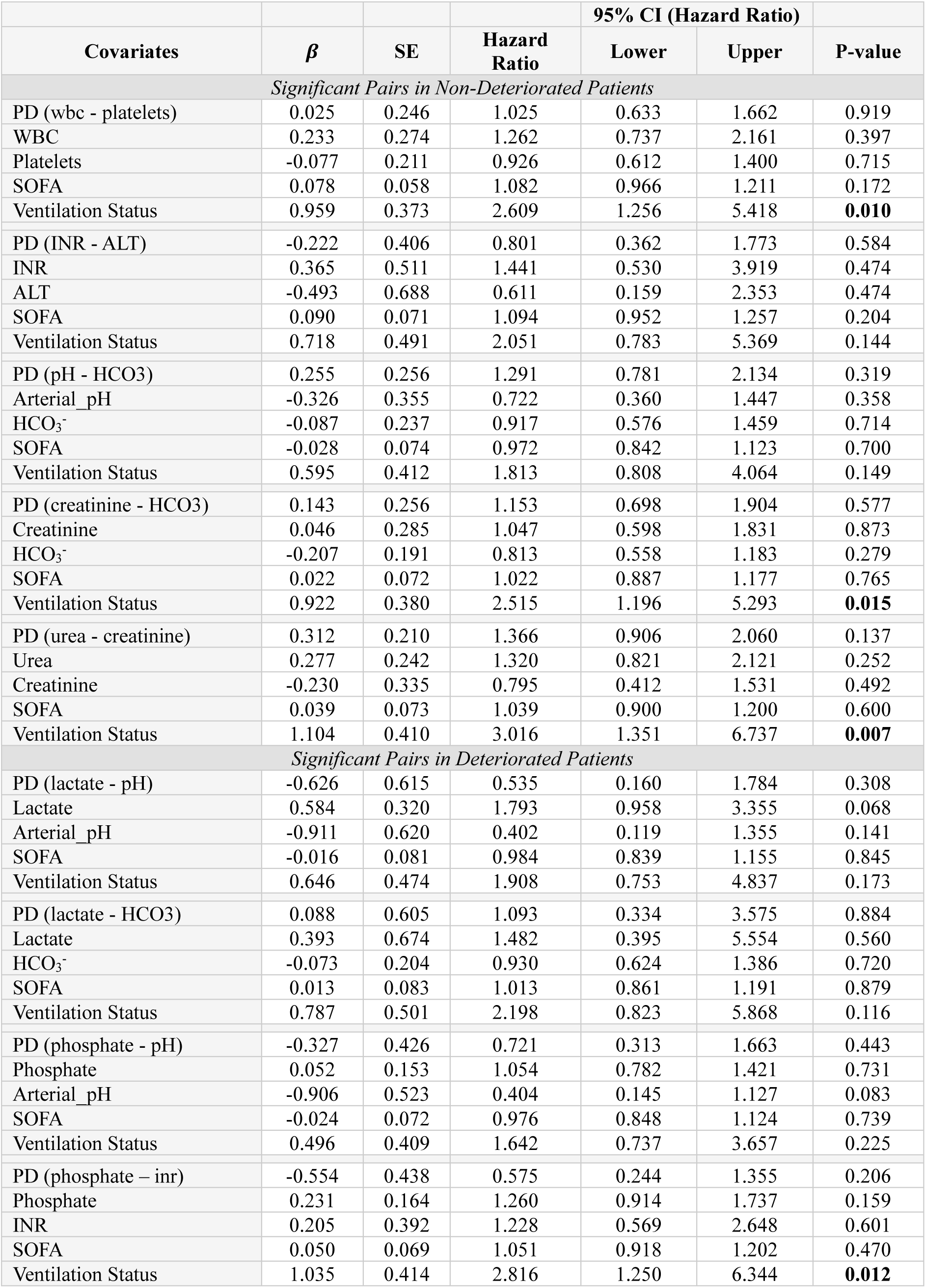
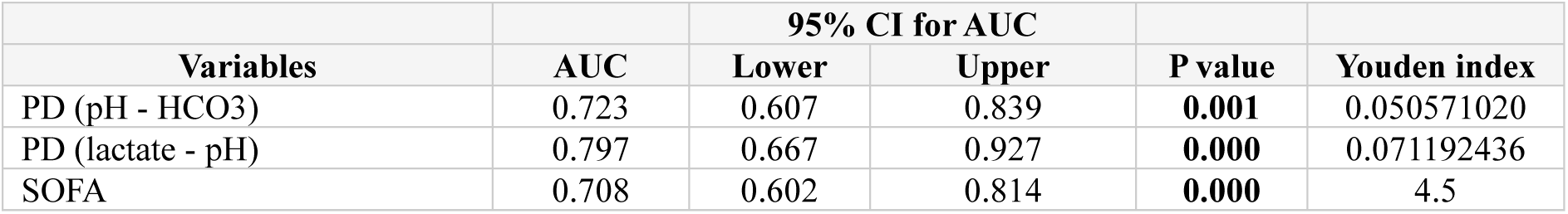
**a.** Demographic and clinical characteristics of the study population with different 30-day survival and 48-hour deterioration outcomes. Data is shown as median (lower quartile – upper quartile) unless stated otherwise as frequency (n) and percentage (%). Chi-squared test was used for evaluating differences in the distribution of positive ventilation, deterioration, mortality, and male sex. Differences in the distribution of other variables were assessed using Mann-Whitney U test. Significant differences in the studied variables (p<0.05) are highlighted in bold. **b.** Mann-Whitney U test results looking at the differences in parenclitic deviations (PD) between survivors “0” and non-survivors “1” for 30-day survival. PD axes significantly different between the two groups are highlighted in bold (p<0.05). **c.** Mann-Whitney U test results looking at the differences in parenclitic deviations (PD) between non-deteriorated “0” and deteriorated “1” patients for 48-hour deterioration. The two groups with significantly different (p < 0.05) PD values are highlighted in bold. **d.** Results from multivariate Cox regression looking at whether parenclitic deviations (PD) can predict 30-day sepsis survival, independent of SOFA and ventilation status. The values of PD in this table were all normalised by z-transformation prior to the analysis. Significant predictors of 30-day mortality (p < 0.05) are highlighted in bold. **e.** Multivariate Cox regression analysis investigating whether parenclitic deviations (PD) can predict 48-hour sepsis deterioration, independent of SOFA and ventilation status. All the PD values in this table were z-normalised prior to the analysis. Significant predictors of 48-hour deterioration (p < 0.05) are highlighted in bold. **f.** Results from ROC Curve Analysis of parenclitic deviations significantly predictive of 30-day mortality (PDs for pH-HCO3 and lactate-pH axes) and SOFA (p<0.05).

| General Characteristics of Survivors and Non-Survivors |  |  |  |
| --- | --- | --- | --- |
|  | Survivors (n=129) | Nonsurvivors (n=33) | P-value |
| Age | 66.3 (51.5-82.3) | 76.1 (67.6-86.1) | **0.008** |
| SOFA score | 4 (2-5) | 6 (3-10) | **<0.001** |
| Male sex, n (%) | 71 (55%) | 23 (69.7%) | 0.128 |
| Positive ventilation status, n (%) | 27 (20.9%) | 18 (54.5%) | **<0.001** |
| Positive 48h deterioration status, n (%) | 8 (6.2%) | 22 (66.7%) | **<0.001** |
| *Physiological variables* |  |  |  |
| Phosphate, mg/dL | 3.1 (2.4-3.7) | 3.9 (2.8-4.4) | **0.004** |
| Arterial pH | 7.39 (7.36-7.43) | 7.32 (7.20-7.42) | **0.024** |
| Urea, mg/dL | 16.5 (10.5-23.3) | 23.3 (15-35.9) | **0.003** |
| Haemoglobin, mg/dL | 11.2 (9.9-12.4) | 11.7 (10.4-12.5) | 0.269 |
| Lactate, mmol/L | 1.6 (1.2-2.4) | 3.8 (1.3-5.9) | **0.002** |
| White blood cell count, $\times 1000/\mu\text{L}$ | 10.8 (8.3-14.1) | 13 (9.8-16.2) | **0.045** |
| Sodium, mEq/L | 139 (136.8-141) | 139 (136.1-143) | 0.738 |
| INR | 1.2 (1.1-1.4) | 1.3 (1.2-1.8) | **0.004** |
| Platelet count, $\times 1000/\mu\text{L}$ | 217 (149-267.5) | 187.3 (138-286.5) | 0.49 |
| Bilirubin, mg/dL | 0.6 (0.4-1.6) | 0.68 (0.43-1.15) | 0.76 |
| Glucose, mg/dL | 124.5 (109.2-150.8) | 143.6 (119.1-178.6) | **0.019** |
| Creatinine, mg/dL | 0.9 (0.7-1.2) | 1.1 (0.9-1.7) | **0.004** |
| Alanine Transaminase, U/L | 28 (17-59.5) | 30 (16.8-149.1) | 0.653 |
| Bicarbonate, mEq/L | 24 (22-26) | 22 (16.6-26.8) | **0.042** |
| Potassium, mEq/L | 4.2 (3.6-4.5) | 3.9 (3.4-5) | 0.967 |
| General Characteristics of Deteriorated and Non-Deteriorated Patients |  |  |  |
|  | No Deterioration (n=132) | Deterioration (n=30) | P value |
| Age | 67.4 (52.1-84.5) | 72.6 (61.8-82.4) | 0.211 |
| SOFA score | 4 (2-6) | 4.5 (3-9.3) | 0.089 |
| Male sex, n (%) | 75 (56.8%) | 19 (63.3%) | 0.514 |
| Positive ventilation status, n (%) | 30 (22.7%) | 15 (50%) | **0.003** |
| Positive 30-day mortality, n (%) | 11 (8.3%) | 22 (73.3%) | **<0.001** |

| *Physiological variables* |  |  |  |
| --- | --- | --- | --- |
| Phosphate, mg/dL | 3.2 (2.4-6.7) | 3.8 (2.6-4.4) | **0.023** |
| Arterial pH | 7.39 (7.36-7.44) | 7.32 (7.19-7.39) | **<0.001** |
| Urea, mg/dL | 16 (10.5-24) | 22.2 (15.5-31.8) | **0.005** |
| Haemoglobin, mg/dL | 11.2 (9.9-12.3) | 11.7 (10.6-12.7) | 0.158 |
| Lactate, mmol/L | 1.6 (1.1-2.4) | 3.7 (2.2-6) | **<0.001** |
| White blood cell count, $\times 1000/\mu\text{L}$ | 10.9 (8.3-14.1) | 11.7 (9.7-16.4) | 0.236 |
| Sodium, mEq/L | 139 (137-141) | 139.5 (135.6-143) | 0.645 |
| INR | 1.2 (1.1-1.4) | 1.3 (1.2-1.5) | 0.145 |
| Platelet count, $\times 1000/\mu\text{L}$ | 217 (147-274) | 187.3 (148.5-282.8) | 0.803 |
| Bilirubin, mg/dL | 0.6 (0.4-1.6) | 0.68 (0.48-0.9) | 0.939 |
| Glucose, mg/dL | 126 (109.8-151.5) | 146.7 (115.9-160.8) | 0.106 |
| Creatinine, mg/dL | 0.9 (0.75-1.2) | 1.1 (0.84-1.7) | **0.028** |
| Alanine Transaminase, U/L | 26 (16-53) | 52.8 (24.1-162.8) | **0.043** |
| Bicarbonate, mEq/L | 24 (22-26) | 22 (16-25.3) | **0.032** |
| Potassium, mEq/L | 4.2 (3.6-4.4) | 3.9 (3.3-4.9) | 0.865 |

|  |  |  |  |  |  |
| --- | --- | --- | --- | --- | --- |
|  |  |  |  |  | **Mann-Whitney U Test** |

|  | 30-day survival | N | Mean | SD | U | r | P-value |
| --- | --- | --- | --- | --- | --- | --- | --- |
| Significant Pairs in Survivors |  |  |  |  |  |  |  |
| PD (WBC - platelets) | 0 | 127 | 3.151 | 3.096 | 2164 | 0.068 | 0.392 |
|  | 1 | 31 | 4.861 | 6.199 |  |  |  |
| PD (INR - ALT) | 0 | 54 | 0.207 | 0.251 | 613 | 0.147 | 0.209 |
|  | 1 | 19 | 0.350 | 0.385 |  |  |  |
| PD (pH - HCO3) | 0 | 64 | 0.045 | 0.034 | 1342 | 0.356 | <0.001 |
|  | 1 | 29 | 0.095 | 0.076 |  |  |  |
| PD (urea - creatinine) | 0 | 127 | 0.219 | 0.174 | 2409 | 0.128 | 0.105 |
|  | 1 | 32 | 0.416 | 0.506 |  |  |  |
| Significant Pairs in Non-Survivors |  |  |  |  |  |  |  |
| PD (WBC - phosphate) | 0 | 119 | 0.658 | 0.454 | 2328 | 0.183 | 0.025 |
|  | 1 | 31 | 1.204 | 1.438 |  |  |  |
| PD (lactate - pH) | 0 | 53 | 0.045 | 0.038 | 844.5 | 0.455 | <0.001 |
|  | 1 | 20 | 0.163 | 0.127 |  |  |  |
| PD (lactate - HCO3) | 0 | 60 | 0.813 | 0.855 | 927.5 | 0.461 | <0.001 |
|  | 1 | 19 | 2.391 | 1.978 |  |  |  |
| PD (urea - creatinine) | 0 | 127 | 0.219 | 0.174 | 2409 | 0.128 | 0.105 |
|  | 1 | 32 | 0.416 | 0.506 |  |  |  |

|  |  |  |  |  | Mann-Whitney U Test |  |  |
| --- | --- | --- | --- | --- | --- | --- | --- |
| PD axis | 48-hour deterioration | N | Mean | SD | U | r | P-value |
| Significant Pairs in Non-Deteriorated Patients |  |  |  |  |  |  |  |
| PD (WBC - platelets) | 0 | 129 | 3.140 | 2.804 | 2001 | 0.047 | 0.558 |
|  | 1 | 29 | 5.130 | 6.825 |  |  |  |
| PD (INR- ALT) | 0 | 56 | 0.248 | 0.293 | 523 | 0.072 | 0.540 |
|  | 1 | 17 | 0.276 | 0.245 |  |  |  |
| PD (pH - HCO3) | 0 | 68 | 0.046 | 0.037 | 1242 | 0.352 | <0.001 |
|  | 1 | 25 | 0.098 | 0.079 |  |  |  |
| PD (creatinine - HCO3) | 0 | 131 | 0.320 | 0.309 | 2585.5 | 0.240 | 0.002 |
|  | 1 | 29 | 0.554 | 0.532 |  |  |  |
| PD (urea - creatinine) | 0 | 131 | 0.213 | 0.173 | 2341.5 | 0.182 | 0.022 |
|  | 1 | 28 | 0.462 | 0.538 |  |  |  |
| Significant Pairs in Deteriorated Patients |  |  |  |  |  |  |  |
| PD (lactate - pH) | 0 | 54 | 0.052 | 0.042 | 678 | 0.243 | 0.038 |
|  | 1 | 19 | 0.141 | 0.130 |  |  |  |
| PD (lactate - HCO3) | 0 | 61 | 0.839 | 0.853 | 817 | 0.352 | 0.002 |
|  | 1 | 18 | 2.364 | 2.027 |  |  |  |
| PD (phosphate - pH) | 0 | 64 | 0.055 | 0.046 | 982.5 | 0.177 | 0.096 |
|  | 1 | 25 | 0.104 | 0.098 |  |  |  |
| PD (phosphate - INR) | 0 | 105 | 0.272 | 0.417 | 1239 | 0.064 | 0.467 |
|  | 1 | 26 | 0.187 | 0.149 |  |  |  |

|  |  |  |  |  |
| --- | --- | --- | --- | --- |
|  |  |  |  | 95% CI (Hazard Ratio) |

| Covariates | $\beta$ | SE | Hazard Ratio | Lower | Upper | P-value |
| --- | --- | --- | --- | --- | --- | --- |
| *Significant Pairs in Survivors* |  |  |  |  |  |  |
| PD (wbc – platelets) | -0.152 | 0.272 | 0.859 | 0.504 | 1.463 | 0.575 |
| WBC | 0.493 | 0.304 | 1.638 | 0.903 | 2.969 | 0.104 |
| Platelets | -0.055 | 0.207 | 0.947 | 0.630 | 1.422 | 0.791 |
| SOFA | 0.215 | 0.054 | 1.240 | 1.115 | 1.378 | **<0.001** |
| Ventilation Status | 1.214 | 0.364 | 3.367 | 1.651 | 6.867 | **<0.001** |
| PD (INR- ALT) | -0.142 | 0.421 | 0.868 | 0.380 | 1.981 | 0.737 |
| INR | 0.665 | 0.557 | 1.945 | 0.653 | 5.792 | 0.232 |
| ALT | -0.801 | 0.850 | 0.449 | 0.085 | 2.378 | 0.346 |
| SOFA | 0.116 | 0.068 | 1.123 | 0.982 | 1.283 | 0.090 |
| Ventilation Status | 1.414 | 0.498 | 4.114 | 1.551 | 10.913 | **0.004** |
| PD (pH - HCO<sub>3</sub>) | 0.733 | 0.217 | 2.081 | 1.361 | 3.184 | **<0.001** |
| Arterial pH | -0.147 | 0.252 | 0.864 | 0.527 | 1.416 | 0.561 |
| HCO<sub>3</sub><sup>-</sup> | -0.160 | 0.205 | 0.852 | 0.571 | 1.273 | 0.435 |
| SOFA | 0.059 | 0.061 | 1.061 | 0.941 | 1.196 | 0.334 |
| Ventilation Status | 0.830 | 0.387 | 2.293 | 1.074 | 4.893 | **0.032** |
| PD (urea - creatinine) | 0.193 | 0.207 | 1.213 | 0.808 | 1.821 | 0.352 |
| Urea | 0.272 | 0.181 | 1.312 | 0.921 | 1.870 | 0.133 |
| Creatinine | -0.059 | 0.313 | 0.943 | 0.510 | 1.743 | 0.851 |
| SOFA | 0.137 | 0.068 | 1.146 | 1.004 | 1.309 | **0.043** |
| Ventilation Status | 1.533 | 0.395 | 4.634 | 2.137 | 10.048 | **<0.001** |
| *Significant Pairs in Non-Survivors* |  |  |  |  |  |  |
| PD (WBC – phosphate) | -0.057 | 0.273 | 0.945 | 0.553 | 1.614 | 0.836 |
| WBC | 0.257 | 0.163 | 1.293 | 0.939 | 1.781 | 0.116 |
| Phosphate | 0.428 | 0.299 | 1.534 | 0.853 | 2.757 | 0.153 |
| SOFA | 0.155 | 0.053 | 1.168 | 1.053 | 1.296 | **0.003** |
| Ventilation Status | 1.229 | 0.372 | 3.418 | 1.648 | 7.089 | **<0.001** |
| PD (lactate - pH) | 1.020 | 0.452 | 2.773 | 1.143 | 6.731 | **0.024** |
| Lactate | 0.137 | 0.275 | 1.147 | 0.668 | 1.968 | 0.619 |
| Arterial pH | -0.042 | 0.359 | 0.959 | 0.474 | 1.939 | 0.908 |
| SOFA | 0.060 | 0.075 | 1.062 | 0.916 | 1.231 | 0.428 |
| Ventilation Status | 0.514 | 0.471 | 1.671 | 0.664 | 4.207 | 0.275 |
| PD (lactate - HCO<sub>3</sub>) | 0.811 | 0.585 | 2.249 | 0.714 | 7.086 | 0.166 |
| Lactate | -0.349 | 0.594 | 0.706 | 0.220 | 2.259 | 0.557 |
| HCO<sub>3</sub><sup>-</sup> | -0.245 | 0.198 | 0.782 | 0.530 | 1.154 | 0.216 |
| SOFA | 0.108 | 0.076 | 1.114 | 0.959 | 1.294 | 0.158 |
| Ventilation Status | 0.985 | 0.510 | 2.678 | 0.985 | 7.276 | 0.053 |
| PD (urea - creatinine) | 0.193 | 0.207 | 1.213 | 0.808 | 1.821 | 0.352 |
| Urea | 0.272 | 0.181 | 1.312 | 0.921 | 1.870 | 0.133 |
| Creatinine | -0.059 | 0.313 | 0.943 | 0.510 | 1.743 | 0.851 |
| SOFA | 0.137 | 0.068 | 1.146 | 1.004 | 1.309 | **0.043** |
| Ventilation Status | 1.533 | 0.395 | 4.634 | 2.137 | 10.048 | **<0.001** |

|  |  |  |  | 95% CI (Hazard Ratio) |  |  |
| --- | --- | --- | --- | --- | --- | --- |
| Covariates | $\beta$ | SE | Hazard Ratio | Lower | Upper | P-value |
| Significant Pairs in Non-Deteriorated Patients |  |  |  |  |  |  |
| PD (wbc - platelets) | 0.025 | 0.246 | 1.025 | 0.633 | 1.662 | 0.919 |
| WBC | 0.233 | 0.274 | 1.262 | 0.737 | 2.161 | 0.397 |
| Platelets | -0.077 | 0.211 | 0.926 | 0.612 | 1.400 | 0.715 |
| SOFA | 0.078 | 0.058 | 1.082 | 0.966 | 1.211 | 0.172 |
| Ventilation Status | 0.959 | 0.373 | 2.609 | 1.256 | 5.418 | 0.010 |
| PD (INR - ALT) | -0.222 | 0.406 | 0.801 | 0.362 | 1.773 | 0.584 |
| INR | 0.365 | 0.511 | 1.441 | 0.530 | 3.919 | 0.474 |
| ALT | -0.493 | 0.688 | 0.611 | 0.159 | 2.353 | 0.474 |
| SOFA | 0.090 | 0.071 | 1.094 | 0.952 | 1.257 | 0.204 |
| Ventilation Status | 0.718 | 0.491 | 2.051 | 0.783 | 5.369 | 0.144 |
| PD (pH - HCO3) | 0.255 | 0.256 | 1.291 | 0.781 | 2.134 | 0.319 |
| Arterial pH | -0.326 | 0.355 | 0.722 | 0.360 | 1.447 | 0.358 |
| HCO<sub>3</sub><sup>-</sup> | -0.087 | 0.237 | 0.917 | 0.576 | 1.459 | 0.714 |
| SOFA | -0.028 | 0.074 | 0.972 | 0.842 | 1.123 | 0.700 |
| Ventilation Status | 0.595 | 0.412 | 1.813 | 0.808 | 4.064 | 0.149 |
| PD (creatinine - HCO3) | 0.143 | 0.256 | 1.153 | 0.698 | 1.904 | 0.577 |
| Creatinine | 0.046 | 0.285 | 1.047 | 0.598 | 1.831 | 0.873 |
| HCO<sub>3</sub><sup>-</sup> | -0.207 | 0.191 | 0.813 | 0.558 | 1.183 | 0.279 |
| SOFA | 0.022 | 0.072 | 1.022 | 0.887 | 1.177 | 0.765 |
| Ventilation Status | 0.922 | 0.380 | 2.515 | 1.196 | 5.293 | 0.015 |
| PD (urea - creatinine) | 0.312 | 0.210 | 1.366 | 0.906 | 2.060 | 0.137 |
| Urea | 0.277 | 0.242 | 1.320 | 0.821 | 2.121 | 0.252 |
| Creatinine | -0.230 | 0.335 | 0.795 | 0.412 | 1.531 | 0.492 |
| SOFA | 0.039 | 0.073 | 1.039 | 0.900 | 1.200 | 0.600 |
| Ventilation Status | 1.104 | 0.410 | 3.016 | 1.351 | 6.737 | 0.007 |
| Significant Pairs in Deteriorated Patients |  |  |  |  |  |  |
| PD (lactate - pH) | -0.626 | 0.615 | 0.535 | 0.160 | 1.784 | 0.308 |
| Lactate | 0.584 | 0.320 | 1.793 | 0.958 | 3.355 | 0.068 |
| Arterial pH | -0.911 | 0.620 | 0.402 | 0.119 | 1.355 | 0.141 |
| SOFA | -0.016 | 0.081 | 0.984 | 0.839 | 1.155 | 0.845 |
| Ventilation Status | 0.646 | 0.474 | 1.908 | 0.753 | 4.837 | 0.173 |
| PD (lactate - HCO3) | 0.088 | 0.605 | 1.093 | 0.334 | 3.575 | 0.884 |
| Lactate | 0.393 | 0.674 | 1.482 | 0.395 | 5.554 | 0.560 |
| HCO<sub>3</sub><sup>-</sup> | -0.073 | 0.204 | 0.930 | 0.624 | 1.386 | 0.720 |
| SOFA | 0.013 | 0.083 | 1.013 | 0.861 | 1.191 | 0.879 |
| Ventilation Status | 0.787 | 0.501 | 2.198 | 0.823 | 5.868 | 0.116 |
| PD (phosphate - pH) | -0.327 | 0.426 | 0.721 | 0.313 | 1.663 | 0.443 |
| Phosphate | 0.052 | 0.153 | 1.054 | 0.782 | 1.421 | 0.731 |
| Arterial pH | -0.906 | 0.523 | 0.404 | 0.145 | 1.127 | 0.083 |
| SOFA | -0.024 | 0.072 | 0.976 | 0.848 | 1.124 | 0.739 |
| Ventilation Status | 0.496 | 0.409 | 1.642 | 0.737 | 3.657 | 0.225 |
| PD (phosphate – inr) | -0.554 | 0.438 | 0.575 | 0.244 | 1.355 | 0.206 |
| Phosphate | 0.231 | 0.164 | 1.260 | 0.914 | 1.737 | 0.159 |
| INR | 0.205 | 0.392 | 1.228 | 0.569 | 2.648 | 0.601 |
| SOFA | 0.050 | 0.069 | 1.051 | 0.918 | 1.202 | 0.470 |
| Ventilation Status | 1.035 | 0.414 | 2.816 | 1.250 | 6.344 | 0.012 |

| Variables | AUC | 95% CI for AUC |  | P value | Youden index |
| --- | --- | --- | --- | --- | --- |
|  |  | Lower | Upper |  |  |
| PD (pH - HCO<sub>3</sub>) | 0.723 | 0.607 | 0.839 | **0.001** | 0.050571020 |
| PD (lactate - pH) | 0.797 | 0.667 | 0.927 | **0.000** | 0.071192436 |
| SOFA | 0.708 | 0.602 | 0.814 | **0.000** | 4.5 |

### Population Correlation Network Maps Identified Distinct Correlation Profiles for 30-day Survival and 48-hour Deterioration

Correlation Network Maps were generated for 162 patients, classified according to 30-day survival and 48-hour deterioration, to explore the population-level physiological network interactions (Figure 3.a.). Four pairs of significantly correlated variables were identified in both survivors and non-survivors for 30-day survival (Bonferroni-corrected p-value≤0.00047619). Similarly, for 48-hour deterioration, four and five pairs of significant physiological correlations were found in non-deteriorated patients and deteriorated patients, respectively. Significant physiological correlations were unique between survivors and non-survivors for 30-day survival and between non-deteriorated and deteriorated patients for 48-hour deterioration, except for the correlation between urea and creatinine which was significant in both survivors and non-survivors. Overall, the correlation profile differed between patients with positive and negative disease outcomes (i.e., survivors versus non-survivors and non-deteriorated versus deteriorated patients); however, it was similar within groups of patients with positive outcomes (i.e., survivors versus non-deteriorated patients) and negative outcomes (i.e., non-survivors versus deteriorated patients) (Figure 3.a.).

**Figure 3.**
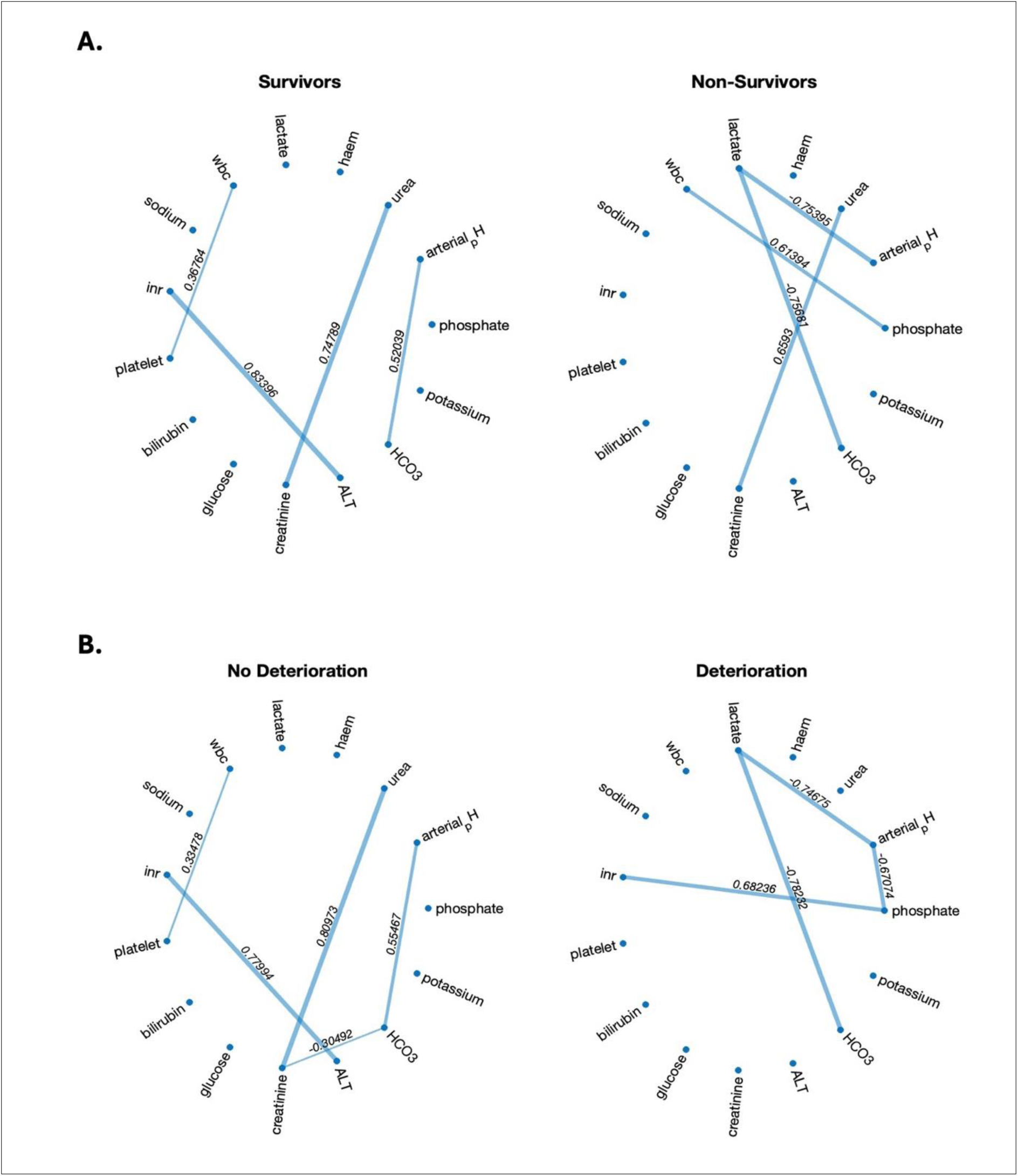
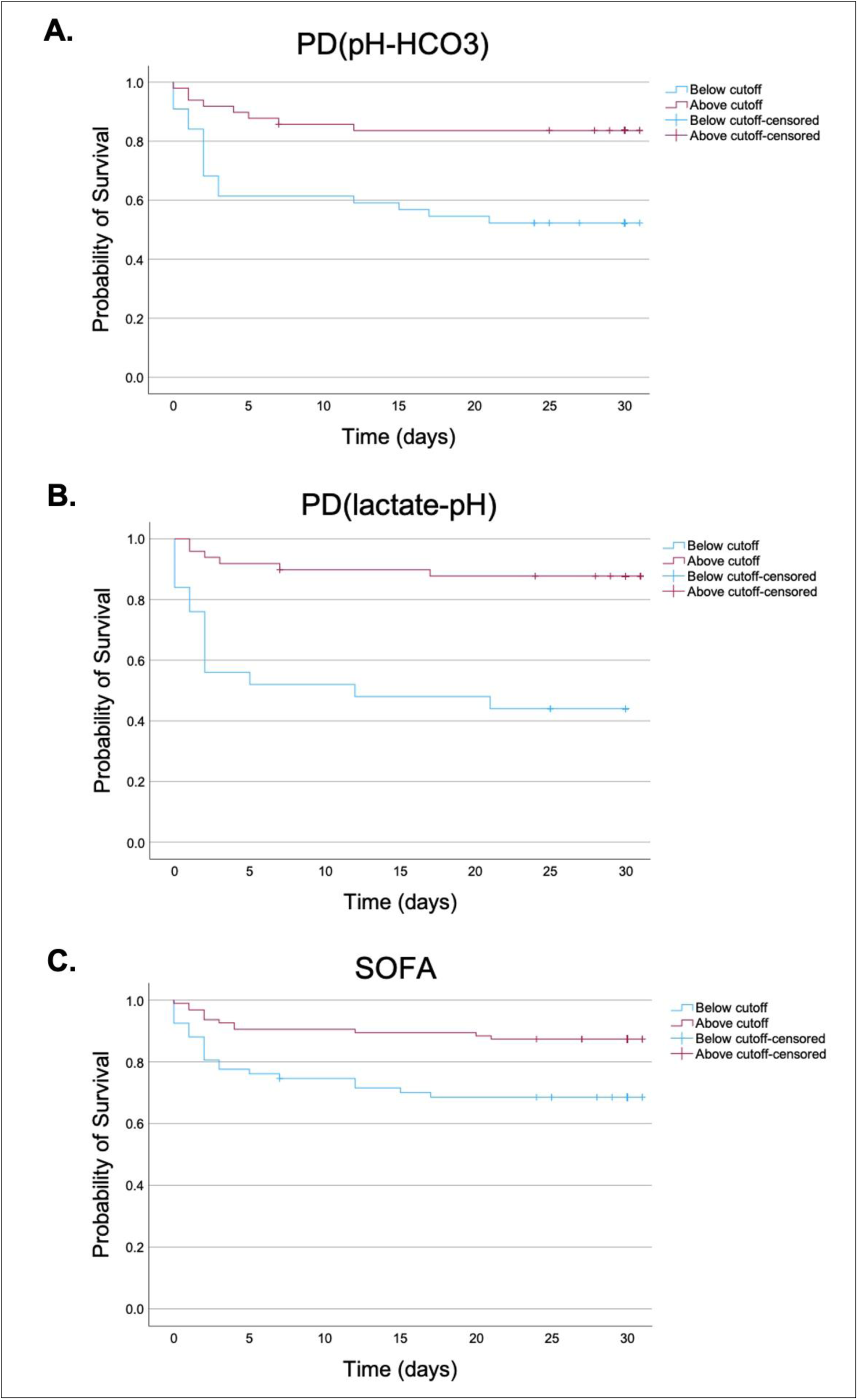
**a.** Population Correlation Network Maps for 30-day survival (A) and 48-hour deterioration (B). Nodes represent physiological variables. Edges show significant correlations (p-value≤Bonferroni-adjusted p-value) between the two nodes. Edge labels display the values of Pearson’s correlation coefficient. **b.** Kaplan-Meier Graphs illustrating 30-day survival of patients classified into predicted survivor and non-survivor groups based on **A**. PD(pH - HCO3), **B**. PD(lactate - pH), and **C**. SOFA score cutoffs. Label in red shows the difference in the survival rates between sepsis patients who were classified above and below the cutoffs.

### Parenclitic Deviations of Significant Physiological Correlations Predict 30-day Mortality Independent of SOFA Score and Ventilation Status but not 48-hour Deterioration in Critically ill Sepsis Patients

Parenclitic deviations (PD) was calculated individually for 162 patients across all significant physiological correlations identified from the population correlation network maps. PDs across the correlated variables were generally higher for non-survivors and deteriorated patients compared to survivors and non-deteriorated patients with small to moderate effect size (r) (Table 3.b. and Table 3.c.) (Cohen, 1988).

Results from the Mann-Whitney U test revealed significant differences in PD between survivors and non-survivors for 30-day survival, along pH-HCO3 axis (p<0.001), WBC-phosphate axis (p=0.025), lactate-pH axis (p<0.001), and lactate-HCO3 axis (p<0.001) (Table 3.b.). Similarly, for 48-hour deterioration, significant differences in PD between patients who did not deteriorate and did deteriorate was found along pH-HCO3 axis (p<0.001), creatinine-HCO3 axis (p=0.002), urea-creatinine axis (p=0.022), lactate-pH axis (p=0.038), and lactate-HCO3 axis (p=0.002) (Table 3.c.).

Subsequently, multivariate Cox regression analysis showed that PDs along pH-HCO3 (hazard ratio, 95% CI = 2.081, 1.361-3.184) and lactate-pH (hazard ratio, 95% CI = 2.773, 1.143-6.731) axes significantly predict 30-day mortality independent of SOFA and ventilation status (Table 3.d.). These results suggest that one standard deviation increase in PD(pH-HCO3) and PD(lactate-pH) can increase the risk of 30-day sepsis mortality by approximately two folds (95% CI= 36% - 3 folds) and three folds (95% CI= 14% - 6 folds), respectively. None of the PDs computed for significant correlation in deteriorated and non-deteriorated patients predicted 48-hour deterioration independent of SOFA and ventilation status (Table 3.e.).

### Cutoffs for Parenclitic Deviations across PD pH-HCO3 and Lactate-pH Axes Significantly Predict 30-day Sepsis Mortality

ROC curve analysis was conducted to evaluate the diagnostic performance of PD(pH-HCO3), PD(lactate-pH) and SOFA score in classifying sepsis patients based on their 30-day survival (p≤0.001). The results indicated that all three measures where moderate classifiers of 30-day sepsis survival. However, the AUC for PDs along pH-HCO3 (AUC, 95%CI = 0.723, 0.607-0.839) and lactate-pH (AUC, 95%CI = 0.797, 0.667-0.927) axes showed a slightly better performance compared to SOFA (AUC, 95%CI = 0.708, 0.602-0.814) with increases in AUC by 2% and 12%, respectively (Table 3.f.).

The Youden Index identified from ROC analysis was used as cut-offs for classifying patients according to their predicted survival status (Table 3.f.). Kaplan-Meier graphs were plotted for PD(pH-HCO3), PD(lactate-pH) and SOFA to compare 30-day survival between the predicted survivors and non-survivors (Figure 3.b.). Log-rank test showed that survival rates were significantly lower in patients classified beyond the cut-offs compared to below the cut-offs for PD(pH-HCO3) (52.3% versus 83.7%, p < 0.001), PD(lactate-pH) (44.0% versus 87.8%, p = 0.001) and SOFA (68.7% versus 87.4%, p = 0.002). The difference in survival rates between patients beyond and below the cut-offs were larger for PDs along pH-HCO3 (Δ = 31.5%) and lactate-pH (Δ = 43.8%) axes compared to SOFA (Δ = 18.7%) (Figure 3.b.).

## Discussion

This report is the first study that investigated physiological network interactions using parenclitic network analysis in both sepsis population and in individual sepsis patients. 162 critically ill sepsis patients were studied retrospectively using health record data from the MIMIC-III database. Fifteen laboratory variables representing different physiological systems were selected to study organ network connectivity in our study population. The main goal of this study was to investigate the physiological network interactions in sepsis patients with varying disease outcomes and to evaluate whether parenclitic network analysis can predict 48-hour deterioration and 30-day sepsis mortality, independent of other markers of sepsis severity. The results indicated that PDs for the pH-bicarbonate axis and pH-lactate axis could significantly predict 30-day mortality in sepsis patients independent of SOFA and ventilation status. None of the PDs were able to predict 48-hour deterioration.

### Correlation Map showed distinct pattern of physiological network interactions between sepsis patients with different disease outcomes

The population correlation analysis was conducted to investigate the pattern of organ network connectivity in patients with different 48-hour deterioration and 30-day survival outcomes. Network correlation maps have shown significant differences in correlation profile between patients with positive and negative disease outcomes (i.e., survivors versus non-survivors). However, the amount of significant physiological correlations (edges) was similar between survivors and non-survivors and between non-deteriorated and deteriorated patients. This indicates that despite the change in the correlation pattern, there was no absolute loss of network connectivity in patients with negative disease outcomes. These findings were different from previous studies investigating organ connectivity in critically ill patients and with patients with chronic liver conditions. A study by Asada et al. investigating critically ill patients from the ICU over a period of 6 months found that non-survivors had considerably less numbers of significant physiological correlations compared to survivors (Asada *et al*., 2016). Similarly, other studies looking at patients with cirrhosis and chronic liver failure, showed that patients who did not survive had a more disrupted organ network (less numbers of edges) compared to those who survived after 3-, 6-, and 12-month follow-up (Tan, Montagnese and Mani, 2020; Zhang *et al*., 2022; Oyelade *et al*., 2023).

However, a recent study looking at the patients with paracetamol-induced acute liver failure (ALF) reported findings that aligned with our results (Oyelade, Moore and Mani, 2024). Population correlation network analysis on ALF patients showed that while there was a general decrease in organ connectivity in non-survivors than the survivors after 28-day follow-up, the scale of disruption was much less severe than in other studies investigating organ connectivity in patients with the chronic liver disease (Tan, Montagnese and Mani, 2020; Zhang *et al*., 2022; Oyelade *et al*., 2023; Oyelade, Moore and Mani, 2024). Similarly to our findings, the study showed that the distinct pattern of organ connectivity between survivors and non-survivors was more evident compared to the presence of physiological network disruption.

A possible explanation for these alignments and discrepancies between the referenced studies and our findings may be due to the variations in the duration of patient follow-up and differences in the nature of the diseases being investigated (i.e., acute versus chronic). Like the study on acute liver failure, which reported results aligning with ours, our study investigated short-term patient mortality by following up the study population for only a month. In contrast, the other studies reporting organ network disruption have investigated patients with chronic conditions (i.e., chronic liver failure) and followed them up over longer period of 3 to 12 months.

Sepsis is a complex and heterogenous, with patients suffering from both short-term and long-term disease outcomes (Delano and Ward, 2016). The absence of disrupted network may be due to difference in physiological response towards acute and chronic conditions. Acute physiological changes in sepsis are often associated with rearrangement of organ network as the body attempts to resist and eliminate infectious pathogens (i.e., via coordination of pro and anti-inflammatory responses and metabolic reprogramming) (Oyelade, Moore and Mani, 2024; Willmann and Moita, 2024). Conversely, chronic changes may exhibit a disrupted organ network connectivity due to development of tolerance (i.e., immunosuppression and chronic inflammation) (Delano and Ward, 2016; Oyelade, Moore and Mani, 2024). Since our study has followed up the patients for only a month, the patients who have passed away are likely to have deceased during the acute stage of disease progression due to organ failure from maladaptive early response towards an attempt to resolve the infection, thus exhibiting distinct pattern of organ connectivity compared to survivors, rather than disruption.

### Acid-base compensatory mechanism and its important role in sepsis survival

Following the population correlation map generation, we investigated organ network connectivity in individual sepsis patients using parenclitic network analysis. Our findings demonstrated that PDs across significant physiological correlations were generally greater in non-survivors and deteriorated patients compared to survivors and non-deteriorated patients, indicating that organ connectivity was more disrupted in patients with negative disease outcomes, while it remained intact in those with positive outcomes. Interestingly, PD values were significantly different between patients with positive and negative disease outcomes across physiological correlations related to metabolic system and acid-base homeostasis; like pH-HCO3, lactate-pH, lactate-HCO3 for both 30-day survival and 48-hour deterioration, and creatinine-HCO3 for 48-hour deterioration only (**Table 3.b.** and **Table 3.c.**).While none of the computed PDs were independent predictive of 48-hour deterioration, results from Cox regression analysis showed that PDs across pH-HCO3 and lactate-pH axes were significantly predictive of 30-day sepsis survival independent of SOFA and ventilation status, suggesting that effective regulation of the acid-base compensatory mechanism could play an important role in sepsis survival.

Cellular metabolism is a widely investigated topic in sepsis research, and metabolic dysfunction in sepsis patients has been reported for almost a century (Thomson, 1929; Liu *et al*., 2022). There is an early shift from oxidative phosphorylation (aerobic respiration) to glycolysis (anaerobic respiration) in immune cells allowing rapid energy production, supporting their activation, proliferation, and differentiation, all of which are essential for generating an effective response to infection during sepsis (Liu *et al*., 2022). However, excessive glycolytic activity and failure to restore oxidative phosphorylation in sepsis have been associated with accumulation of lactate (a by-product of anaerobic respiration) and reduction in blood pH (Suetrong and Walley, 2016). Under normal physiological conditions, a reduction in blood pH is countered by bicarbonate buffering, returning pH to its physiological range. However, in sepsis, persistent lactatemia due to excessive glycolysis can overwhelm the bicarbonate buffer system, depleting the plasma bicarbonate levels (Kamel, Oh and Halperin, 2020). This dysregulation of the acid-base homeostasis can lead to metabolic acidosis and contribute to organ dysfunction.

The underlying mechanisms behind dysfunction in the cellular metabolism and the acid-base compensatory system in sepsis remains an active area of research. Numerous studies investigating skeletal muscle and liver samples in sepsis have shown that reduction in mitochondrial activity is linked to poor disease outcomes (Brealey *et al*., 2002; Haden *et al*., 2007; Carré *et al*., 2010). In particular, nitric oxide (NO) is overproduced in sepsis due to inflammatory stress and is known to influence the mitochondrial respiration (Brown, 1999; Cauwels, 2007). An animal study investigating the effects of NO on mitochondrial activity has shown that high levels of NO can inhibit mitochondrial complexes I, II, II, and IV by competing with oxygen, thereby reducing aerobic respiration. While the inhibition of complex II, III, and IV were reversible as NO levels steadily decline, prolonged high exposure to NO can irreversibly inhibit complex I and lead to cytotoxic effects (Clementi *et al*., 1998). Other studies investigating muscle biopsies from septic patients also demonstrated significantly elevated NO production, reduced mitochondrial complex I activity, lower ATP production, and elevated serum lactate levels in non-survivors compared to survivors (Brealey *et al*., 2002; Carré *et al*., 2010). These results support that persistent NO inhibition of mitochondrial enzymes can result in permanent mitochondrial damage. Additionally, provide strong evidence that inability to restore oxidative metabolism may lead to a cascade event (i.e., lactate overproduction due to increased anaerobic respiration, metabolic acidosis, and organ dysfunction) resulting in poor sepsis outcomes (Suetrong and Walley, 2016). Consequently, increase in PD along pH-lactate may be due to physiological uncoupling in the metabolic system, particularly in the homeostasis between glycolysis and oxidative phosphorylation.

Nonetheless, metabolic dysfunction and lactate overproduction are not the only factors that may contribute to the dysregulation of acid-base balance in sepsis non-survivors. Other processes, such as reduced bicarbonate production also is likely to be involved. Under healthy conditions, loss of endogenous bicarbonate is primarily compensated by the liver through the Cori cycle (Cori and Cori, 1929). Excess lactate produced from anaerobic respiration in peripheral tissue is transported to the liver and converted back to glucose via gluconeogenesis. During this process, bicarbonate is produced on a 1:1 basis as a by-product of lactate metabolism, which helps to correct the acid-base imbalance (Heering *et al*., 1999; Katopodis, Pappas and Katopodis, 2022). However, in sepsis, this lactate-mediated bicarbonate generation may not be happening fast enough to compensate for the elevated lactate production. Two studies investigating liver glucose production in septic rats showed that gluconeogenesis from lactate was significantly reduced in septic rats compared to the healthy controls (Clemens *et al*., 1983; de Souza Galia *et al*., 2021). Potential reasons for reduced lactate-driven gluconeogenesis in sepsis have been reported by several studies. Firstly, impaired gluconeogenesis could be due to reduced activity of essential enzymes involved in gluconeogenesis (i.e., pyruvate carboxylase, phosphoenolpyruvate carboxy kinase) (Deutschman *et al*., 1993; Wang *et al*., 2015).

Secondly, impaired mitochondrial respiration and reduced ATP production in sepsis could potentially slow down gluconeogenesis, as this process is energy demanding (Rognstad, 1982; de Souza Galia *et al*., 2021). Therefore, increase in PD along pH-bicarbonate axis in non-survivors, could be due to dysregulation within the acid-base compensatory system, particularly in the balance between lactate production and lactate clearance via gluconeogenesis.

Since, PDs across pH-HCO3 and lactate-pH axes predicted sepsis survival, we also looked at how the original levels of lactate, HCO3, and pH varied between patients with different disease outcomes. Analysis of general clinical characteristics showed that patients with poor disease outcomes (non-survivors and deteriorated patients) had a significantly higher lactate, lower bicarbonate, and decreased pH compared to patients with positive disease outcomes (survivors and non-deteriorated patients) (Table 3.a.). These findings further support that sepsis patient with poor disease outcomes exhibit a dysregulated pattern of the acid-base homeostasis. Similarly, a prospective study investigating acidosis patterns in sepsis patients revealed a distinct pattern of change in levels of blood lactate, bicarbonate, and pH between survivors and non-survivors (Ganesh *et al*., 2016). Ganesh et al. showed that on the day of ICU admission, both survivors and non-survivor groups had a higher lactate (>2mmol/L), lower HCO3 (<23.0mEq/L) and decreased pH (<7.39), all of which were outside the normal healthy range (Jones, 2010; Ganesh *et al*., 2016; Raphael *et al*., 2016). However, when examining the temporal change in these parameters over the period of 5 days, the study showed that the lactate levels decreased and bicarbonate and pH levels increased, approaching normal range in survivors. In contrast, in non-survivors, the lactate levels remained persistently high, and the bicarbonate and pH levels continued to stay low (Ganesh *et al*., 2016). These results suggest that there is difference in the acute metabolic and acid-base compensatory responses between sepsis survivors and non-survivors, supporting our earlier finding, where we identified a distinct pattern of organ network connectivity between patients with different disease outcomes.

Overall, these observations support the critical roles of cellular metabolism and acid-base compensatory mechanism in shaping the early course of sepsis, which may either lead to an adaptive response that restores body homeostasis, or a maladaptive response causing physiological dysregulation. Furthermore, these findings highlight that parenclitic analysis can not only predict sepsis survival but can also provide useful information about sepsis pathophysiology and difference in physiological response between survivors and non-survivors.

### Conventional (i.e., SOFA) versus Network Physiological approaches (i.e., PD and TE) for sepsis evaluation

One of our main goals was to assess whether network physiological techniques, such as parenclitic deviation, could offer a more effective evaluation of early sepsis outcomes compared to conventional approach like SOFA. Our findings showed that both SOFA and PDs for pH-HCO3 and lactate-pH axes were predictive of 30-day survival but not 48-hour deterioration (Table 3.d. and Table 3.e.). Hence, we performed ROC curve analysis to compare the diagnostic performances between parenclitic network mapping and SOFA at predicting sepsis survival.

Our results show that the AUC for PDs across pH-HCO3 (0.723) and lactate-pH (0.797) axes were higher compared to SOFA (0.708), indicating that PD had a better specificity and specificity at predicting 30-day sepsis survival compared to SOFA. We also plotted Kaplan-Meier graphs for PDs and SOFA, using the Youden Index from the ROC curve analysis as a cut-off for classifying patients into predicted survivors and non-survivor groups (Figure 3.b.). The Kaplan-Meier graphs showed that the difference in survival rates between patients above and below the cut-off for PDs along pH-HCO3 (Δ = 31.5%) and lactate-pH (Δ = 43.8%) axes was around two folds larger than the difference observed for SOFA (Δ = 18.7%). This finding suggests that the cut-offs for PDs have a stronger ability to differentiate survivors and non-survivors than SOFA, supporting the idea that network physiological approaches like parenclitic network mapping perform better at evaluating sepsis survival compared to the other linear techniques.

Earlier in the introduction, we have mentioned that there are at least two different techniques for studying organ network interactions within individual patients. The first technique known as dynamic network mapping (i.e., transfer entropy) analyses time-series data such as dynamical changes of vital sign recordings (heart rate, respiratory rate and oxygen saturation). The second method, called parenclitic network mapping (i.e. a static network mapping) computes cross-sectional data like blood test results. The dynamic network mapping was previously investigated in sepsis by Morandotti et al. The present study was a continuation of this study and investigated the application of static network mapping in predicting sepsis outcomes, using the same sepsis population included in the earlier work by Morandotti et al. (Morandotti *et al*., 2025).

We were interested in comparing the effectiveness between dynamic (transfer entropy) and static (parenclitic deviation) network mapping techniques in predicting sepsis outcomes. Hence, we compared our findings with the work by Morandotti et al. which looked at information transfer between the cardiorespiratory parameters - heart rate (HR), respiratory rate (RR), and blood oxygen saturation (SpO_2_) - using transfer entropy and evaluated whether it could predict early sepsis deterioration and survival (Morandotti *et al*., 2025). Similarly to our findings with PD, Morandotti et al. found that the transfer entropy (TE) values, representing the amount of information transferred between SpO2 → HR, HR → RR, and RR → HR were significantly predictive of 30-day sepsis survival, independent of other markers of sepsis severity (Supplementary Figure S2). Consequently, we compared the diagnostic performance between TE and PD using results from the ROC curve analysis. We found that AUCs for TEs (0.635-0.724) were comparable to that of SOFA (0.708), while the AUCs for PDs (0.723-0.797) were generally higher than those for TEs (Supplementary Table S1). This result suggests that PDs are better than TEs at predicting 30-day sepsis survival, as they demonstrate higher specificity and sensitivity for distinguishing potential sepsis survivors from non-survivors. However, unlike PD or SOFA, TEs were significantly predictive of 48-hour deterioration, independent of other factors impacting sepsis outcomes (Supplementary Figure S3).

Collectively, this information offers several useful clinical insights. The fact that PD has a better predictive performance at classifying 30-day sepsis survival, implies that in resource-limited settings where obtaining clean time-series data is challenging, PD analysis using laboratory data can supplement SOFA to improve identification of patients at risk of having an early sepsis death. On the other hand, the inability for the parenclitic analysis to predict 48-hour deterioration unlike transfer entropy, indicates that static network approach using point-in-time data could potentially overlook subtle physiological changes happening over time, which is otherwise detected in the dynamic network mapping.

Network physiological approaches including dynamic network mapping studied by Morandotti et al. (2025) and static network mapping investigated in the present work offered independent information to SOFA and performed better at predicting sepsis outcomes. This supports the limitations of linear model of sepsis (i.e., SOFA) in evaluating sepsis and highlights the potential role of network physiological approaches in improving early detection of sepsis outcomes.

### Future Perspective

Parenclitic deviation (PD) not only predicts early sepsis mortality but also provides valuable insight into individual patients’ disease pathogenesis. Employing PD in ICU settings may improve understanding of distinct biological phenotypes within sepsis population and potentially facilitate the development of personalised treatments based on physiological network map generated for each patient (Singer, 2019; Iglesias *et al*., 2020; Oyelade *et al*., 2023).

### Limitations

Parenclitic network mapping is a promising approach for evaluating sepsis outcomes, however several limitations need to be considered. The sample size of our study cohort (n=162) was small, due to the strict exclusion criteria requiring both clean timeseries and laboratory data. This limitation affects the statistical power of our results. Moreover, this paper was a retrospective study that investigated patient data from the MIMIC-III database, which was collected from a single hospital in North America. This study design made our study population relatively homogenous, limiting the generalisability of our results to wider sepsis population across different regions. Future studies should validate our findings by using a larger, multi-center cohort, to evaluate applicability of parenclitic network mapping across heterogenous sepsis population with diverse clinical and demographic backgrounds.

There were further limitations within our statistical analysis. For population-level correlation analysis, we used parametric correlation analysis, in the upcoming studies, using other non-parametric alternatives would be more generalizable, as it does not hold any assumptions about the underlying data distribution. Subsequently, for the multivariate Cox regression, the covariates were only limited to SOFA and ventilation status. While these two are common markers of sepsis severity, other variables such as pre-existing comorbidities and use of medications (i.e., beta-blockers) are also known to influence sepsis outcomes (Chu *et al*., 2024; Kang *et al*., 2024). Therefore, future studies should investigate whether prognostic ability of PD is influenced by these additional factors.

### Conclusion

Parenclitic deviation predicted 30-day mortality in sepsis patients but not 48-hour deterioration, independent of other markers of sepsis severity. This method may offer useful insight into sepsis pathophysiology and different physiological response between sepsis patients with various disease outcomes. Parenclitic network mapping analyses routine laboratory data and is a promising tool for early identification and treatment of at-risk sepsis sub-population in the clinical settings. Further research is required to validate our findings in a larger multi-centered sepsis cohort.

## Data availability

Data will be made available upon reasonable request.

## Supplemental material

Supplemental material Figure S1: https://doi.org/10.6084/m9.figshare.28218341.v1

Supplemental material Figure S2: https://doi.org/10.6084/m9.figshare.28218401.v1

Supplemental material Figure S3: https://doi.org/10.6084/m9.figshare.28218359.v1

Supplemental material Table S1: https://doi.org/10.6084/m9.figshare.28218410.v1

## Acknowledgements

The authors are grateful to Dr Anika Cawthorn (UCL Advanced Research Computing Centre) for collaboration and support.

## Author contributions

E.I., T.O., M.W., W.L., and A.R.M. conceived and designed research; E.I., M.W., J.L., and T.O., analysed data; E.I., and A.R.M. interpreted results of experiments; E.I. prepared figures; E.I. drafted manuscript; T.O., J.L., and A.R.M. edited and revised manuscript; E.I., T.O., M.W., J.L., W.L., and A.R.M. approved final version of manuscript.

